# Safety and immunogenicity of a variant-adapted SARS-CoV-2 recombinant protein vaccine with AS03 adjuvant as a booster in adults primed with authorized vaccines

**DOI:** 10.1101/2022.12.02.22282931

**Authors:** Guy de Bruyn, Joyce Wang, Annie Purvis, Martin Sanchez Ruiz, Haritha Adhikarla, Saad Alvi, Matthew I Bonaparte, Daniel Brune, Agustin Bueso, Richard M Canter, Maria Angeles Ceregido, Sachin Deshmukh, David Diemert, Adam Finn, Remi Forrat, Bo Fu, Julie Gallais, Paul Griffin, Marie-Helene Grillet, Owen Haney, Jeffrey A Henderson, Marguerite Koutsoukos, Odile Launay, Federico Martinon Torres, Roger Masotti, Nelson L Michael, Juliana Park, Doris M Rivera M, Natalya Romanyak, Chris Rook, Lode Schuerman, Lawrence D Sher, Fernanda Tavares-Da-Silva, Ashley Whittington, Roman M Chicz, Sanjay Gurunathan, Stephen Savarino, Saranya Sridhar, the VAT00002 booster cohorts study team

## Abstract

**Background:** Booster vaccines providing protection against emergent SARS-CoV-2 variants are needed. In an international phase 3 study, we evaluated booster vaccines containing prototype (D614) and/or Beta (B.1.351) variant recombinant spike proteins and AS03 adjuvant (CoV2 preS dTM-AS03).

**Methods:** Adults, primed 4–10 months earlier with mRNA (BNT162b2, mRNA-1273]), adenovirus-vectored (Ad26.CoV2.S, ChAdOx1nCoV-19) or adjuvanted protein (CoV2 preS dTM-AS03 [D614]) vaccines and stratified by age (18-55 and ≥56 years), were boosted with monovalent (MV) D614 (5μg, n=1285), MV (B.1351) (5μg, n=707) or bivalent (BiV) (2.5μg D614 plus 2.5μg B.1.351, n=625) CoV2 preS dTM-AS03. SARS-CoV-2-naïve adults (controls, n=479) received a primary series (two injections, 21 days apart) of CoV2 preS dTM-AS03 containing 10μg D614. Antibodies to D614G, B.1.351 and Omicron BA.2 and BA.1 variants were evaluated using validated pseudovirus (lentivirus) neutralization (PsVN) assay. D614G or B.1.351 PsVN titers 14 days (D15) post-booster were compared with pre-booster (D1) titers in BNT162b2-primed participants (18-55 years old) and controls (D36), for each booster formulation (co-primary objectives). Safety was evaluated throughout the trial. Results of a planned interim analysis are presented.

**Results:** Among BNT162b2-primed adults (18-55 years old), PsVN titers against D614G or B.1.351 were significantly higher post-booster than anti-D614G titers post-primary vaccination in controls, for all booster formulations, with an anti-D614G GMT ratio (98.3% CI) of 2.16 (1.69; 2.75) for MV(D614), an anti-B.1.351 ratio of 1.96 (1.54; 2.50) for MV (B.1.351) and anti-D614G and anti-B.1.351 ratios of 2.34 (1.84; 2.96) and 1.39 (1.09; 1.77), respectively, for BiV. All booster formulations elicited cross-neutralizing antibodies against Omicron BA.2 across vaccine priming subgroups and against Omicron BA.1 (evaluated in BNT162b2-primed participants). Similar patterns in antibody responses were observed for participants aged ≥56 years. No safety concerns were identified.

**Conclusion:** CoV2 preS dTM-AS03 boosters demonstrated acceptable safety and elicited robust neutralizing antibodies against multiple variants, regardless of priming vaccine.

**ClinicalTrials.gov:** NCT04762680

**Funding:** Sanofi and federal funds from the Biomedical Advanced Research and Development Authority (BARDA), part of the office of the Administration for Strategic Preparedness and Response at the U.S. Department of Health and Human Services under Contract # HHSO100201600005I, and in collaboration with the U.S. Department of Defense Joint Program Executive Office for Chemical, Biological, Radiological and Nuclear Defense under Contract # W15QKN-16-9-1002.

## Introduction

The COVID-19 pandemic has seen an unprecedented deployment of COVID-19 vaccines.^1-3^ The continued emergence of new variants of concern (VoC) of the severe acute respiratory syndrome coronavirus 2 (SARS-CoV-2), together with waning immunity and protection against mild and moderately symptomatic infection, underscore the need for new and updated booster vaccines to enhance and broaden protection.^4-9^ Bivalent boosters, targeting the prototype virus and Omicron BA.1 or BA.4 and BA.5 subvariants have recently been authorized for use in the EU, USA and the UK.^10-13^ While Omicron variant-containing vaccines were designed to include the predominant circulating strain, an alternative approach is to include strains that provide broad cross-protection against emergent variants.^14^

Sanofi and GSK have developed a SARS-CoV-2 recombinant protein vaccine with AS03 adjuvant (CoV2 preS dTM-AS03) using a baculovirus vector system to express stabilized SARS-CoV-2 pre-fusion S protein (preS dTM).^15^ In parallel with development of a D614 strain containing booster and primary series vaccine^16^, we developed and are evaluating a CoV2 preS dTM-AS03 booster formulation containing 5 μg of B.1.351 variant S protein (monovalent B.1.351) and a formulation containing 2.5 μg of D614 plus 2.5 μg of B.1.351 variants (bivalent D614+B.1.351). In previously primed non-human primates, Beta (B.1.351) variant-containing adjuvanted recombinant protein vaccines showed broad cross-reactivity across SARS-CoV-2 variants of concern and SARS-CoV-1.^17,18^

Here, we present interim safety and immunogenicity data for three CoV2 preS dTM-AS03 booster formulations (monovalent D614, monovalent B.1.351 and bivalent D614+B.1.351) in participants from our phase 3 cohorts, who were previously primed with authorized COVID-19 mRNA or adenovirus-vectored vaccines, or with CoV2 preS dTM-AS03 (5, 10 or 15 μg of D614 antigen).

## Methods

### Study design and participants

In a substantial amendment to the Phase 2 primary series study (NCT04762680)^16^ on June 10 2021, two supplemental cohorts (cohort 1 and cohort 2) were added for phase 3 evaluation of the safety and immunogenicity of monovalent D614, monovalent B.1.351 and bivalent D614+B.1.351 CoV2 preS dTM-AS03 booster formulations (**Figure S1**). Booster groups in both cohorts included adults (≥18 years) from France, the USA, the UK, Australia and Spain who had completed a primary series of an authorized or approved D614 mRNA vaccine (2 doses of BNT162b2 [30 μg; Pfizer/BioNTech] or mRNA-1273 [100 μg; Moderna]) or adenovirus-vectored vaccine (2 doses of ChAdOx1 nCoV-19 [5×10^10^ vp; Oxford University/AstraZeneca] or 1 dose of Ad26.CoV2.S [5×10^10^ vp; J&J/Janssen]) at least 4 months and no longer than 10 months prior to enrolment. Cohort 2 included a subgroup of adults from the USA and Honduras who were previously primed in the original phase 2 study 4–10 months earlier with two doses of CoV2 preS dTM-AS03 containing 5, 10 or 15 μg of D614 antigen. Cohort 1 included a parallel, non-randomized control group, encompassing unvaccinated adults, 18– 55 years of age, from the USA and Australia who tested negative for antibodies to SARS-CoV-2 with a rapid diagnostic test (COVID-19 IgG and IgM Rapid Test Cassette; Healgen Scientific, Houston, TX, USA). Individuals with pre-existing medical conditions, those who were immunocompromised (except those with organ transplant in the past 180 days, chemotherapy in the past 90 days or with HIV and CD4 counts <200/mm^3^) and those with a potentially increased risk for severe COVID-19^19^ were eligible for participation in the study. The exclusion criteria are described in full in the **Supplementary Appendix, Methods**. Vaccine administration was open-label in cohort 1 booster and control groups and modified double-blind (observer-blinded) in cohort 2.

Participants were enrolled between 29 July 2021 and 22 October 2021 to cohort 1 and between 15 November 2021 and 22 February 2022 to cohort 2. Here, we report interim immunogenicity data up to 14 days after last vaccination in all participants and safety data up to cut-off dates 18 February 2022 for cohort 1 and 13 May 2022 for cohort 2. For controls, long-term safety analyses were conducted up to 13 May 2022.

The study was undertaken in compliance with the International Conference on Harmonization guidelines for Good Clinical Practice and the principles of the Declaration of Helsinki. The protocol and amendments were approved by Independent Ethics Committees (country ethics committees for Honduras [Comité Ética Independiente Zugueme], France [Comite De Protection Des Personnes Ile De France III Hôpital Tarnier-Cochin, France], Spain [Comité De Éticade Investigación Con Medicamentos Parc Taulï, Spain] and the UK [HRA and Health and Care Research Wales, UK], a central ethics committee for the USA [Advarra] and local ethics committees for the USA [WCG IRB, Columbia Research Human Research Protection Office, Langone Health Office of Science and Research Institutional Review Board and the Yale Human Research Protection Program] and for Australia [Bellberry, Sydney Children’s Hospitals Network Human Research Ethics Committee and Sydney Children’s Hospital Network (SCHN) Research Governance]) as per local regulations. Written informed consent was obtained from all participants before any study procedures were performed.

### Procedures

Booster cohort participants were stratified by priming vaccine and by age category (18–55 years or ≥56 years) (**Figure S1**). In the cohort 1 booster groups, all participants were offered a dose of monovalent D614. In cohort 2, participants primed with an mRNA or adenovirus-vectored vaccine were randomized 1:1 to receive monovalent B.1.351 or bivalent D614+B.1.351 boosters; while those previously primed with CoV2 preS dTM-AS03 (5, 10 or 15 μg of D614 antigen) were randomized 9:1 (18–55-year-olds) or 1:1 (≥56 year-olds) to receive monovalent D614 or monovalent B.1.351 boosters, respectively. Participants in the control group received two injections, 21 days apart (D1 and D22), of CoV2 preS dTM-AS03 containing 10 μg of D614 antigen.

The CoV2 preS dTM-AS03 vaccine was described previously.^15,16^ Preparation of the booster formulations and intramuscular administration are described in **Supplementary Methods**.

### Immunogenicity

SARS-CoV-2 neutralizing antibody responses were measured using a lentivirus-based pseudovirus neutralization (PsVN) assay expressing the full-length S protein of the SARS-CoV-2 D614G, Beta (B.1.351) or Omicron (BA.1 or BA.2) variants (Monogram Biosciences LabCorp, South San Francisco, CA, USA; **Supplementary Methods**).^20^

Primary endpoints were individual serum PsVN titers at D1 and D15 against D614G for monovalent D614 recipients, against B.1.351 for monovalent B.1.351 recipients, against D614G and B.1.351 for bivalent D614+B.1.351 recipients, and against D614G 14 days after the last dose (D36) for the control group. Secondary endpoints included PsVN titers against D614G and B.1.351 for each predefined timepoint, the percentage of participants reaching a ≥4-fold rise in serum neutralization titer post-vaccination (D15 for booster cohorts; D22 and D36 for the control group) relative to pre-vaccination (D1) and individual serum neutralization titer fold-rise. Exploratory endpoints included individual serum PsVN titers against Omicron subvariants BA.2 and BA.1 in available samples across all priming vaccine subgroups for BA.2 and from BNT162b2-primed participants for BA.1.

### Safety

Safety was assessed for each treatment group and by age category (18–55 years or ≥56 years). Safety endpoints were as described in the original phase 2 study,^16^ with the list of AEs of special interest (AESIs) updated to include anaphylactic reactions, generalized convulsions, thrombocytopenia, thrombosis with thrombocytopenia syndrome, myocarditis, pericarditis and potential immune-mediated diseases (pIMDs).^21,22^ Adverse events (AEs) were graded from grade 1 (mild) to grade 3 (severe; prevents normal daily activities) and assessed by the investigator as related or unrelated to the study vaccine.

### Statistics

Co-primary immunogenicity objectives were to demonstrate, for BNT162b2-primed participants 18– 55 years of age: i) the non-inferiority of post-booster D614G PsVN responses following monovalent D614, post-booster B.1.351 PsVN responses following monovalent B.1.351 or both responses following bivalent D614+B.1.351, compared to the D614G PsVN response elicited by primary vaccination in the control group; and ii) the superiority of the post-booster relative to pre-booster PsVN response. Statistical inference was based on the use of two-sided 98.3% CIs. Non-inferiority in terms of neutralizing antibody titers was concluded if the lower bound of the 98.3% CI for the between-group (booster versus control) GMT ratio was >0.67. Superiority in terms of neutralizing antibody titers was concluded if the lower limit of the 2-sided 98.3% CI of the geometric mean of individual ratio (GMTR; post-booster versus pre-booster) was >2. Study objectives are described in more detail in the **Supplementary Methods**.

The per-protocol analysis set (PPAS) comprised all participants who received the booster injection or both primary vaccinations (control group), who met all protocol inclusion and no exclusion criteria, provided blood samples on D15 (booster cohorts) or D22 and D36 (control group), did not have pre-specified protocol deviations and did not receive a COVID-19 vaccine outside of this study before D15 (booster cohorts) or D36 (control group). Analyses in the control group were performed on SARS-CoV-2 naïve participants from the PPAS. Naïve or non-naïve status was determined based on serological detection of anti-S antibodies on D1 (S-ELECSYS; Elecsys Anti-SARS-CoV-2 S, Roche, Indianapolis, IN, USA EUA202698) and nucleic-acid amplification test (NAAT; Abbott RealTime SARS-CoV-2 assay, EUA200023) for detection of SARS-CoV-2 in nasopharyngeal swabs on D1 and D22 (**Supplementary Methods**). The safety analysis set (SafAS) comprised all participants who received at least one study vaccine dose, with analysis according to the vaccine actually received.

For full details on statistical considerations, including planned sample sizes, see **Supplementary Methods**. Statistical analyses were performed using SAS® Version 9.4 or later.

## Results

### Participants and participant disposition

Allocated booster doses were administered to 803 of 806 participants enrolled into the monovalent D614 group, 705 of 707 participants enrolled into the monovalent B.1.351 group and 621 of 625 enrolled into the bivalent D614+B.1.351 group. In the control group, 473 of 479 enrolled SARS-CoV-2-naïve participants received at least one primary dose of monovalent D614. A total of 18 monovalent D614 booster recipients, 34 monovalent B.1.351 booster recipients, 26 bivalent D614+B.1.351 recipients and 43 participants in the control group discontinued the study before the analysis cut-off (**Figure S2**). The median interval between last primary and booster doses tended to be slightly shorter overall for monovalent D614 recipients (5.75 months) than for monovalent B.1.351 or bivalent D614+B.1.351 recipients (6.96 and 6.50 months, respectively) (**Table 1**). Across the three booster groups (SafAS), the mean age (standard deviation [SD]) ranged from 43.7 (14.3) to 50.4 (15.0) years; 22.2%–37.5% were ≥56 years old. Control group participants had a mean age of 37.5 (11.2) years. The majority of participants (65.5%–78.7% across booster and control groups) were white. Across booster and control groups, 50.8%–59.8% of participants had at least one high-risk medical condition (**Table 1**), of which obesity was the most common (**Table S1**). Among booster groups in the PPAS, prior SARS-CoV-2 infection based on anti-nucleoprotein seropositivity was detected at baseline for 68/734 (9.3%) of monovalent D614 recipients, 138/615 (22.4%) of monovalent B.1.351 recipients and 124/561 (22.1%) of bivalent vaccine recipients.

**Table 1:**
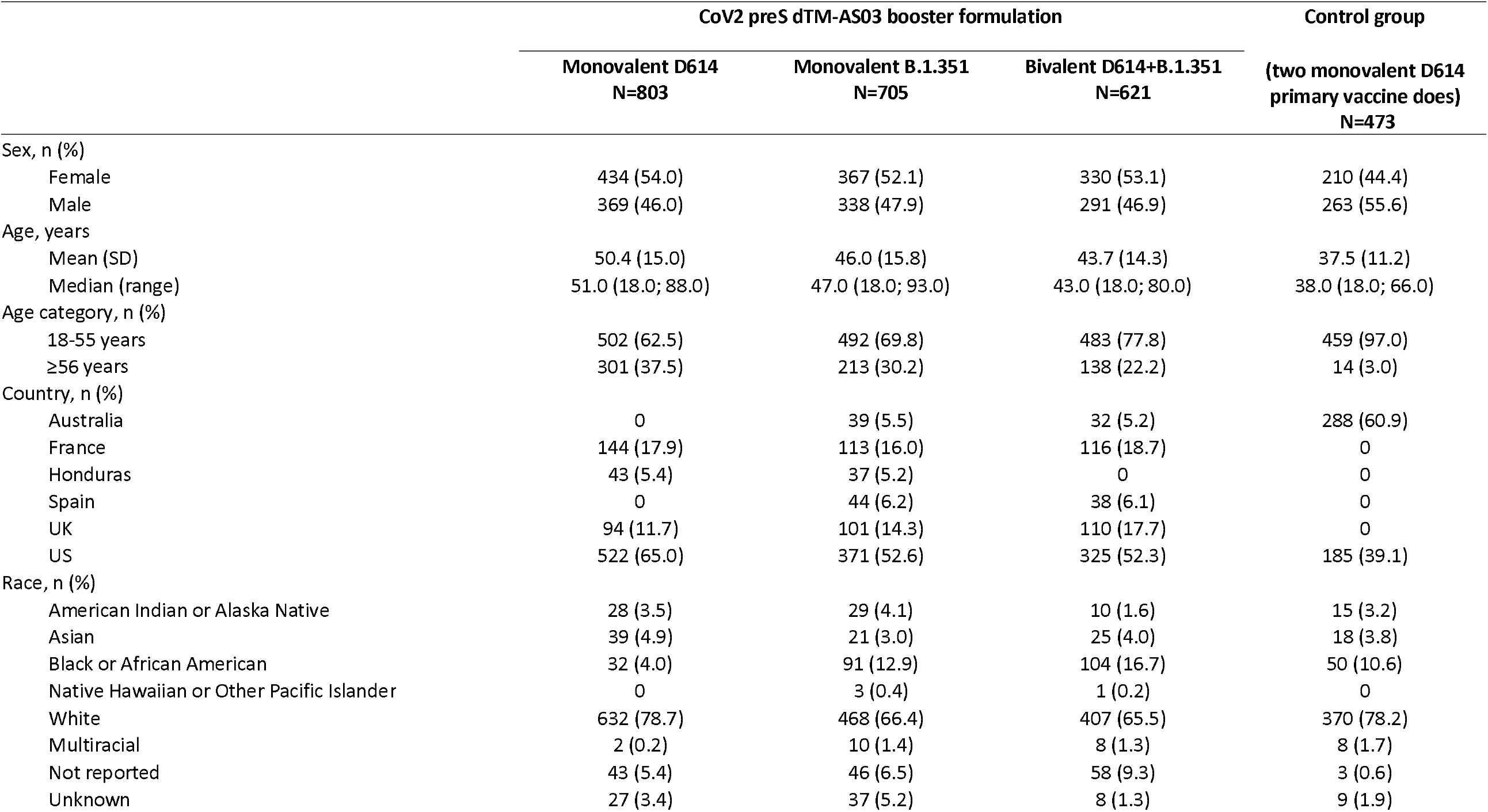
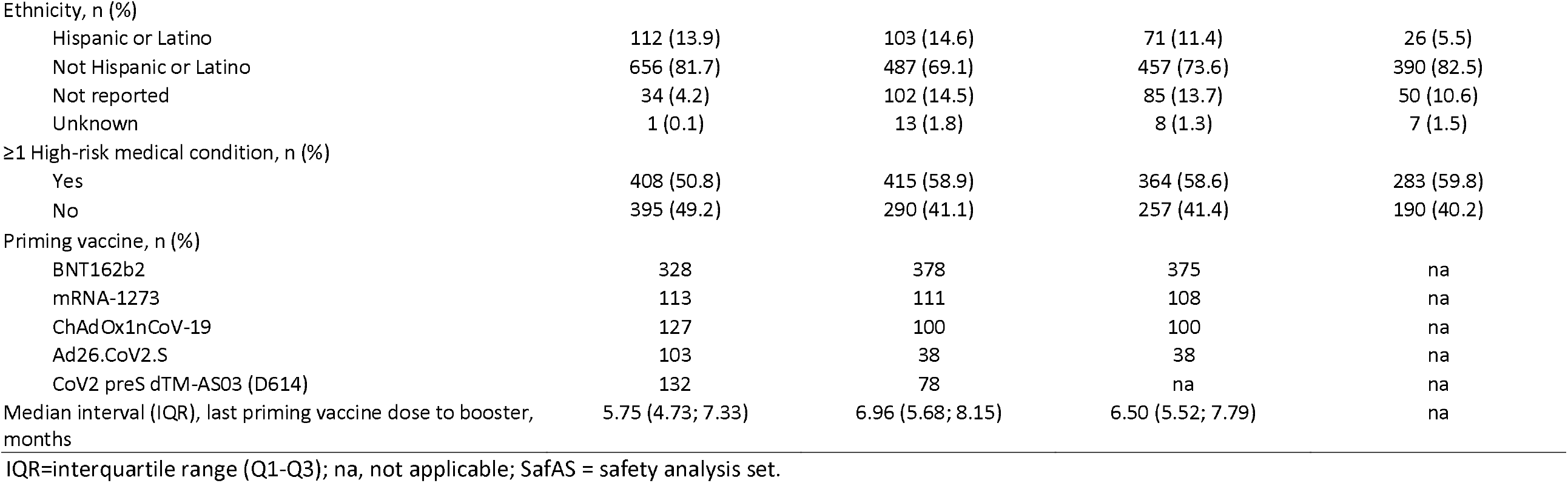
Participant demographic characteristics (SafAS)

|  | CoV2 preS dTM-AS03 booster formulation |  |  | Control group |
| --- | --- | --- | --- | --- |
|  | Monovalent D614<br>N=803 | Monovalent B.1.351<br>N=705 | Bivalent D614+B.1.351<br>N=621 | (two monovalent D614<br>primary vaccine doses)<br>N=473 |
| Sex, n (%) |  |  |  |  |
| Female | 434 (54.0) | 367 (52.1) | 330 (53.1) | 210 (44.4) |
| Male | 369 (46.0) | 338 (47.9) | 291 (46.9) | 263 (55.6) |
| Age, years |  |  |  |  |
| Mean (SD) | 50.4 (15.0) | 46.0 (15.8) | 43.7 (14.3) | 37.5 (11.2) |
| Median (range) | 51.0 (18.0; 88.0) | 47.0 (18.0; 93.0) | 43.0 (18.0; 80.0) | 38.0 (18.0; 66.0) |
| Age category, n (%) |  |  |  |  |
| 18-55 years | 502 (62.5) | 492 (69.8) | 483 (77.8) | 459 (97.0) |
| ≥56 years | 301 (37.5) | 213 (30.2) | 138 (22.2) | 14 (3.0) |
| Country, n (%) |  |  |  |  |
| Australia | 0 | 39 (5.5) | 32 (5.2) | 288 (60.9) |
| France | 144 (17.9) | 113 (16.0) | 116 (18.7) | 0 |
| Honduras | 43 (5.4) | 37 (5.2) | 0 | 0 |
| Spain | 0 | 44 (6.2) | 38 (6.1) | 0 |
| UK | 94 (11.7) | 101 (14.3) | 110 (17.7) | 0 |
| US | 522 (65.0) | 371 (52.6) | 325 (52.3) | 185 (39.1) |
| Race, n (%) |  |  |  |  |
| American Indian or Alaska Native | 28 (3.5) | 29 (4.1) | 10 (1.6) | 15 (3.2) |
| Asian | 39 (4.9) | 21 (3.0) | 25 (4.0) | 18 (3.8) |
| Black or African American | 32 (4.0) | 91 (12.9) | 104 (16.7) | 50 (10.6) |
| Native Hawaiian or Other Pacific Islander | 0 | 3 (0.4) | 1 (0.2) | 0 |
| White | 632 (78.7) | 468 (66.4) | 407 (65.5) | 370 (78.2) |
| Multiracial | 2 (0.2) | 10 (1.4) | 8 (1.3) | 8 (1.7) |
| Not reported | 43 (5.4) | 46 (6.5) | 58 (9.3) | 3 (0.6) |
| Unknown | 27 (3.4) | 37 (5.2) | 8 (1.3) | 9 (1.9) |
| Ethnicity, n (%) |  |  |  |  |
| Hispanic or Latino | 112 (13.9) | 103 (14.6) | 71 (11.4) | 26 (5.5) |
| Not Hispanic or Latino | 656 (81.7) | 487 (69.1) | 457 (73.6) | 390 (82.5) |
| Not reported | 34 (4.2) | 102 (14.5) | 85 (13.7) | 50 (10.6) |
| Unknown | 1 (0.1) | 13 (1.8) | 8 (1.3) | 7 (1.5) |
| ≥1 High-risk medical condition, n (%) |  |  |  |  |
| Yes | 408 (50.8) | 415 (58.9) | 364 (58.6) | 283 (59.8) |
| No | 395 (49.2) | 290 (41.1) | 257 (41.4) | 190 (40.2) |
| Priming vaccine, n (%) |  |  |  |  |
| BNT162b2 | 328 | 378 | 375 | na |
| mRNA-1273 | 113 | 111 | 108 | na |
| ChAdOx1nCoV-19 | 127 | 100 | 100 | na |
| Ad26.CoV2.S | 103 | 38 | 38 | na |
| CoV2 preS dTM-AS03 (D614) | 132 | 78 | na | na |
| Median interval (IQR), last priming vaccine dose to booster, months | 5.75 (4.73; 7.33) | 6.96 (5.68; 8.15) | 6.50 (5.52; 7.79) | na |

IQR=interquartile range (Q1-Q3); na, not applicable; SafAS = safety analysis set.

### Immunogenicity

#### Monovalent D614 booster

Among participants aged 18–55 years, the D614G PsVN GMT for BNT162b2-primed participants increased from 339 at D1 to 7894 (95% CI: 6993; 8911) on D15 following monovalent D614 booster, demonstrating superiority of the PsVN response post-booster compared to pre-booster (post/pre-booster ratio, 23.37 [98.3% CI: 18.58; 29.38]). PsVN titers against D614G at D15 were twice the level of those induced after primary vaccination with CoV2 preS dTM-AS03 (D614) in the control group (D36 GMT: 3658 [95% CI 3123; 4286]) demonstrating non-inferiority (GMT ratio of 2.16 [98.3% CI: 1.69; 2.75]) to the control group. Thus, the co-primary objectives for the monovalent D614 booster group were met (**Table 2A;** see **Table S3** for PsVN titers in the control group).

**Table 2.** Non-inferiority and superiority of PsVN titers against D614G and B.1.351 following monovalent and bivalent boosters compared with CoV2 preS dTM-AS03 (D614) primary vaccination or compared with pre-booster PsVN titers - PPAS

**A. Monovalent D614 booster group**
| Priming vaccine or vaccine platform subgroup | Booster D15 |  | Control D36 |  | Post-booster vs control |  |  | Post-booster vs pre-booster |  |  |  |
| --- | --- | --- | --- | --- | --- | --- | --- | --- | --- | --- | --- |
|  | Strain | M | Strain | M | GMT ratio | 98.3% CI | NI* | M | GMTR | 98.3% CI | Superiority† |
| BNT162b2 | D614G | 202 | D614G | 302 | 2.16 | 1.69; 2.75 | Yes | 189 | 23.37 | 18.58; 29.38 | Yes |
| mRNA vaccine platform | D614G | 269 | D614G | 302 | 2.09 | 1.66; 2.64 | Yes | 255 | 18.08 | 14.75; 22.16 | Yes |
| Pooled Ad virus vectored platform | D614G | 131 | D614G | 302 | 1.89 | 1.43; 2.49 | Yes | 108 | 30.18 | 20.56; 44.31 | Yes |
| CoV2 preS dTM-AS03 (D614) | D614G | 52 | D614G | 302 | 7.27 | 5.34; 9.89 | Yes | 45 | 51.63 | 22.29; 119.57 | Yes |

**B. Monovalent B.1.351 booster group**
| Priming vaccine or vaccine platform subgroup | Booster D15 |  | Control D36 |  | Post-booster vs control |  |  | Post-booster vs pre-booster |  |  |  |
| --- | --- | --- | --- | --- | --- | --- | --- | --- | --- | --- | --- |
|  | Strain | M | Strain | M | GMT ratio | 98.3% CI | NI* | M | GMTR | 98.3% CI | Superiority† |
| BNT162b2 | B.1.351 | 279 | D614G | 302 | 1.96 | 1.54; 2.50 | Yes | 256 | 35.41 | 26.71; 46.95 | Yes |
|  | B.1.351 | 279 | B.1.351 | 291 | 17.36 | 13.39; 22.50 | Yes‡ | - | - | - | - |
| mRNA vaccine platform | B.1.351 | 347 | D614G | 302 | 2.06 | 1.62 ; 2.61 | Yes | 321 | 34.19 | 26.58 ; 43.98 | Yes |

**C. Bivalent D614+B.1.351 booster group**
| Priming vaccine or vaccine platform subgroup | Booster D15 |  | Control D36 |  | Post-booster vs control |  |  | Post-booster vs pre-booster |  |  |  |
| --- | --- | --- | --- | --- | --- | --- | --- | --- | --- | --- | --- |
|  | Strain | M | Strain | M | GMT ratio | 98.3% CI | NI* | M | GMTR | 98.3% CI | Superiority† |
| BNT162b2 | D614G | 276 | D614G | 302 | 2.34 | 1.84; 2.96 | Yes | 269 | 14.39 | 11.34; 18.28 | Yes |
|  | B.1.351 | 276 | D614G | 302 | 1.39 | 1.09; 1.77 | Yes | 248 | 34.18 | 25.84; 45.22 | Yes |
|  | B.1.351 | 276 | B.1.351 | 291 | 12.31 | 9.50; 15.97 | Yes‡ | - | - | - | - |
| mRNA vaccine platform | D614G | 343 | D614G | 302 | 2.52 | 2.00 ; 3.16 | Yes | 332 | 13.04 | 10.56 ; 16.11 | Yes |
|  | B.1.351 | 342 | D614G | 302 | 1.47 | 1.16 ; 1.85 | Yes | 309 | 31.19 | 24.33 ; 39.97 | Yes |

D, study day; M, number of participants with available data for the analysis; NI, non-inferiority
\*Non-inferiority of a booster response against D614G or B.1.351 compared to a response against D614G in the control group was concluded if the lower bound of the 98.3% CI for the between-group (booster versus control) GMT ratio was $>0.67$ .
†Superiority of a booster response compared to a pre-booster response was concluded if the lower limit of the 2-sided 98.3% CI of the fold-rise (post-booster versus pre-booster) was $>2$ .
‡Superiority, rather than non-inferiority, was demonstrated for this comparison (conditional secondary objective). The superiority of a booster dose response against B.1.351 of monovalent B.1.351 or bivalent D614+B.1.351 in BNT162b2 primed participants aged 18-55 years compared to a response against B.1.351 in the control group was concluded if the lower limit of the 2-sided 98.3% CI of the ratio of GMTs between groups was $> 1.5$ .

Non-inferiority versus the control group was demonstrated for the pooled mRNA vaccine-primed group (GMT ratio, 2.09 [98.3% CI: 1.66; 2.64]), the pooled adenovirus vectored vaccine-primed group (GMT ratio, 1.89 [98.3% CI: 1.43; 2.49]) and the CoV2 preS dTM-AS03 (D614)-primed group (GMT ratio, 7.27 [98.3% CI: 5.34; 9.89]). Superiority versus the pre-booster response was also demonstrated for the mRNA-primed group (GMTR, 18.08 [98.3% CI: 14.75; 22.16]), for the adenovirus-vectored primed group (GMTR, 30.18 [98.3% CI: 20.56; 44.31]) and the CoV2 preS dTM-AS03 (D614) group (GMTR, 51.63 [98.3% CI: 22.29; 119.57]) (**Table 2A**). The magnitude of D614G PsVN titers at D15 following monovalent D614 booster in 18–55 year olds were similar in the mRNA and adenoviral vectored priming groups but higher in the group primed with the CoV2 preS dTM-AS03 (D614) vaccine (**Figure 1A; Table S2**). Seroresponse rates to D614G ranged from 71.2% to 93.1% across priming vaccine subgroups following monovalent D614 booster and was 99.0% (on D36) in the control group among 18–55 year olds (**Table S2**).

**Figure 1.**
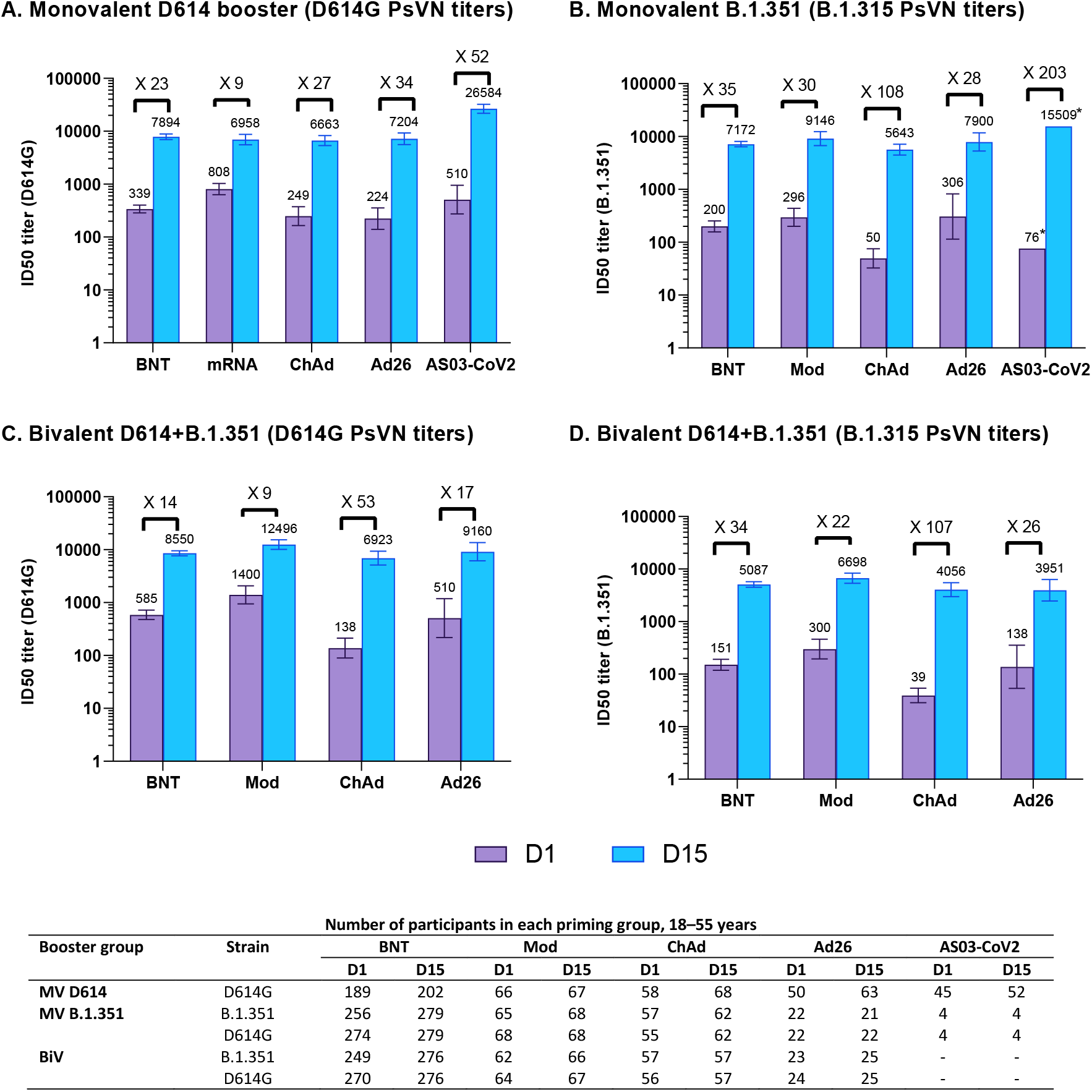
Booster pseudovirus neutralizing antibody responses against D614G or B.1.351 strains for (A) monovalent D614, (B) monovalent B.1.351 and (C and D) bivalent D614+B.1.351 among 18–55 year olds, by prior priming vaccine – PPAS. Graphs are annotated with geometric mean titers (above each bar) and geometric means of individual titer ratios (above grouped bars), calculated based on paired post-vaccination/pre-vaccination results. Error bars denote 95% CIs for the GMTs. *95% confidence not calculated due to small number of participants analyzed (n=4). Table shows number of participants with available data for at each timepoint. D1, study day 1 (pre-booster); D15, study day 15 (14 days post-booster); BNT, BNT162b2-primed; Mod, mRNA-1273-primed; ChAd, ChAdOx1nCoV-19-primed; Ad26, Ad26.CoV2.S-primed; AS03-CoV2, CoV2 preS dTM-AS03 (D614)-primed

A similar pattern in D614G PsVN titers was observed in older adults (≥56 years) (**Figure S3**). While post-booster GMTs in older adults tended to be of a lower magnitude than those in younger adults, they were still numerically higher than D36 GMTs post-primary vaccination in the control group, for all vaccine-primed subgroups (**Table S2**).

#### Monovalent B.1.351 booster

In BNT162b2-primed participants aged 18–55 years, B.1.351 PsVN GMTs increased from 200 at D1 to 7172 (95% CI 6363; 8083) at D15 following a booster dose of monovalent B.1.351 (**Figure 1B**) demonstrating superiority to pre-booster titers (D15/D1 titer ratio 35.41 [98.3% CI 26.71; 46.95]) (**Table 2B**). Comparing B.1.351 PsVN titers following the booster to those following primary vaccination demonstrated non-inferiority to D614G PsVN antibody GMTs (GMT ratio, 1.96 [98.3% CI 1.54; 2.50]) and superiority to B.1.351 (GMTR, 17.36 [98.3% CI 13.39; 22.50]) (**Table 2B**).

Among participants aged 18–55 years, B.1.351 PsVN GMTs (95% CI) following monovalent B.1.351 booster increased from between 49.7 (32.7; 75.6) and 306 (114; 822) at D1 to between 5643 (4468; 7128) and 9146 (6734; 12421) at D15 across the mRNA and adenovirus-vectored priming vaccine subgroups (**Figure 1B**). D15 GMTs were 1.5- to 2.5-fold higher than D614G PsVN GMTs following primary vaccination in the control group. PsVN titers (95% CI) against D614G also increased in the monovalent B.1.351 booster group (18–55 years age category) across the four mRNA or adenovirus-vectored priming vaccine subgroups from between 165 (105; 258) and 1331 (917; 1931) at D1 to between 6817 (5453; 8521) and 13189 (9836; 17684) at D15. Seroresponse rates to B.1.351 ranged from 71.4% to 96.5% and those to D614G, from 63.6%–89.1% across priming vaccine subgroups (**Table S4**).

PsVN GMTs against B.1.351 and D614G following monovalent B.1.351 booster also increased among older adults (≥ 56 years), with D15 GMTs in all vaccine priming subgroups exceeding those following primary vaccination in the control group (**Figure S3; Table S4**). D15 GMTs in older adults tended to be lower than those in younger participants in all but the mRNA-1273-primed group. Notably, the increase in post-booster PsVN titers following monovalent B.1.351 booster was particularly marked for CoV2 preS dTM-AS03 (D614)-primed participants (≥56 years; N=68), with PsVN GMTs (95% CI) rising to 24407 (17705; 33647) (anti-D614G) and 13180 (9571; 18151) (anti-B.1.351). This marked increase in titers was also seen in the 18–55 years age category for a small number (n=4) of participants (**Table S4**).

#### Bivalent (D614+B.1.351) booster

Among BNT162b2-primed participants aged 18–55 years, PsVN GMTs (95% CI) to D614G increased from 585 (477; 718) at D1 to 8550 (7638; 9571) at D15 and PsVN GMTs to B.1.351 increased from 151 (119; 191) at D1 to 5087 (4511; 5737) at D15 following a bivalent booster dose, demonstrating superiority of D614G and B.1.351 booster responses at D15 compared to pre-booster (D15/D1 GMTRs [98.3% CI]: 14.39 [11.34; 18.28] and 34.18 [25.84; 45.22], respectively) (**Figure 1C-D; Table 2C**). The non-inferiority of PsVN titers against D614G and B.1.351 following bivalent booster compared to post-primary D614G PsVN titers in the control group was also demonstrated, with GMT ratios (98.3% CI) of 2.34 (1.84; 2.96) and 1.39 (1.09; 1.77), respectively. In addition, B.1.351 PsVN titers induced by the bivalent booster were demonstrated to be superior to B.1.351 PsVN titers in the control group (GMT ratio, 12.31 [98.3% CI: 9.50; 15.97]) (**Table 2C**).

Among 18–55 year olds, D614G and B.1.351 PsVN GMTs following the bivalent booster increased substantially across all vaccine priming subgroups, with GMTRs (post-/pre-booster) ranging from 9 to 53 for anti-D614G and from 21 to 107 for anti-B.1.351 (**Figure 1C-D**). Seroresponse rates to D614G ranged from 61.9% (48.8; 73.9) to 89.1% (77.8; 95.9) and those to B.1.351 ranged from 78.3 (95% CI: 56.3; 92.5) to 96.4% (95% CI: 87.7; 99.6) across subgroups (**Table S5**).

Increases in neutralizing antibody titers (against B.1.351 and against D614G) following the bivalent D614+B.1.351 booster were also seen among older adults (≥56 years) (D15 GMTs: 2025–10035 [B.1.351]; 4492–22531 [D614G] across subgroups), with similar or lower PsVN titers compared to younger adults in all but the Ad26.CoV2.S-primed subgroup, in which GMTs were higher in older than younger adults (**Table S5**).

#### Neutralizing antibody responses to Omicron subvariant strains (exploratory analysis)

In a subset of participants aged 18–55 years (N=132 in the monovalent D614 booster; N=160 in the monovalent B.1.351 group; N=156 in the bivalent booster group), baseline BA.2 PsVN titers were higher in the monovalent B.1.351 and bivalent groups than in the monovalent D614 group. Omicron BA.2 PsVN GMTs increased 19 to 25-fold from baseline following booster, across the three booster groups (**Figure 2A)** with the highest titers induced by the monovalent B.1.351 booster (2663 [95% CI: 2260; 3137]), followed by the bivalent booster (1810 [95% CI: 1577; 2077]) and monovalent D614 booster (953 [95% CI: 797; 1141]). In older adults (≧56 years) (N=74 in the monovalent D614 booster; N=59 in the monovalent B.1.351 group; N=33 in the bivalent booster group), while baseline titers were comparable across booster groups, higher titers were elicited by the monovalent B.1.351 than by the other formulations. Similar results were obtained for booster responses against the Omicron BA.1 subvariant in BNT162b2-primed participants (**Figure 2B**).

**Figure 2.**
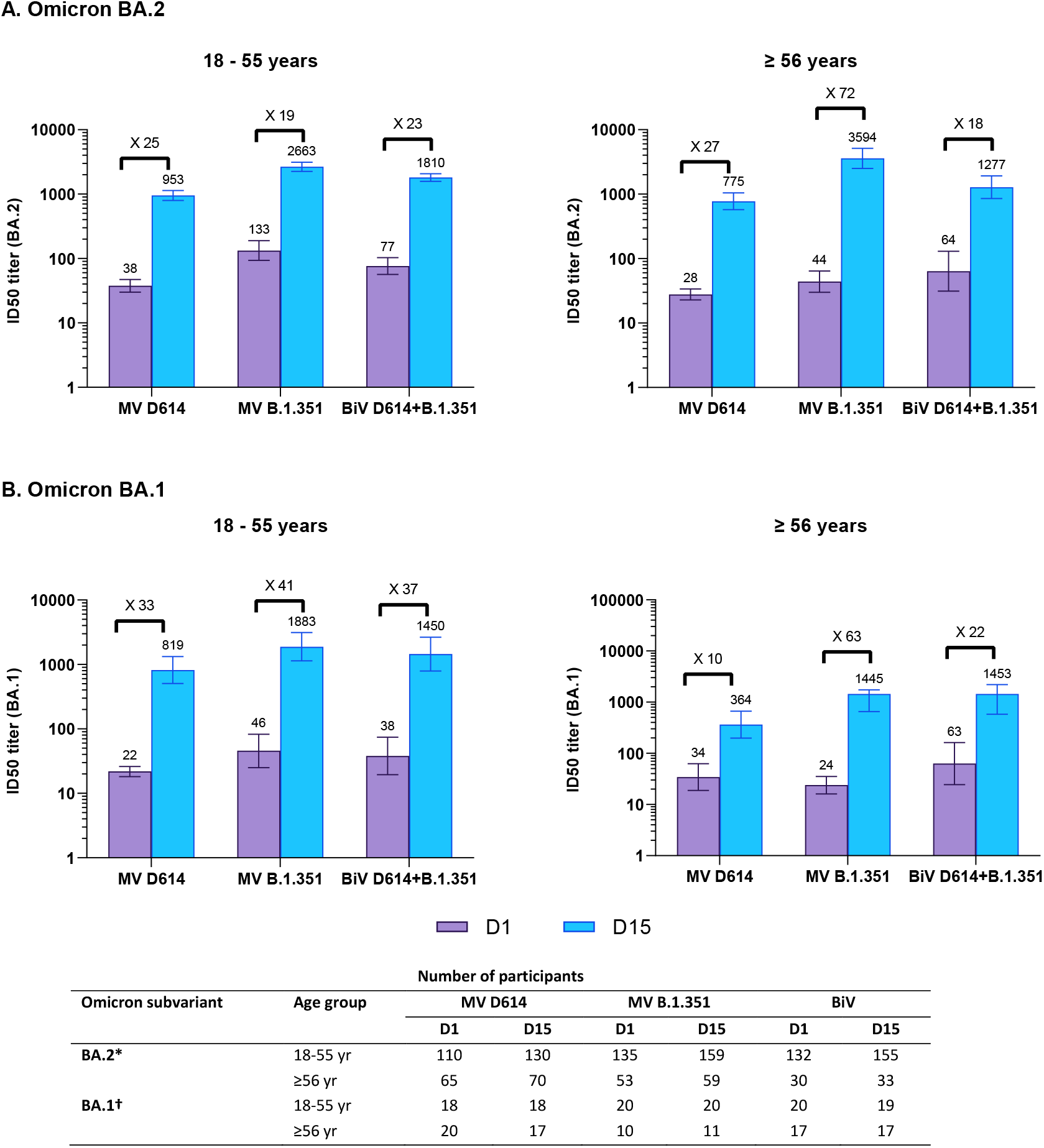
Cross-neutralizing titers elicited by monovalent and bivalent CoV2 preS dTM-AS03 formulations against (A) Omicron BA.2 and (B) BA.1, by age category. MV, monovalent; BiV, bivalent Graphs are annotated with geometric mean titers (above each bar) and geometric means of individual titer ratios (above grouped bars), calculated based on paired post-vaccination/pre-vaccination results. Error bars denote 95% CIs for the GMTs, calculated using normal approximation of log-transformed titers. Table shows number of participants with available data for at each timepoint. *Assessed on available samples across priming vaccine subgroups; †Assessed on available samples in the BNT162b2-primed subgroup

### Safety

In each booster group, ≥98.0% of participants had completed at least 2 months of safety follow-up by the analysis cut-off dates; the median duration of safety follow-up was 144–150 days across booster groups. In the control group, 70.9% had completed ≥2 months safety follow-up and the median duration was 148 days.

Adverse events for each booster formulation are summarized in **Table S6**. There were no immediate unsolicited AEs reported in any booster group. Within 7 days after booster, grade 3 solicited injection reactions were reported by 2.5% participants in the monovalent D614 booster group, 3.3% in the monovalent B.1.351 group and 1.9% in the bivalent group (**Figure 3**); grade 3 solicited systemic reactions were reported by 6.6%, 6.9% and 6.3%, respectively (**Figure 4**). Most solicited reactions were transient (mostly lasting 1–3 days) and self-limited; all participants fully recovered. Within 21 days after booster, 7.1–8.8% participants reported at least one unsolicited adverse reaction. Across priming vaccine subgroups, solicited systemic reactions tended to be more frequently reported for CoV2 preS dTM-AS03 (D614)-primed participants than for those primed with other priming vaccines (**Figure S4**). Solicited and unsolicited events were mostly reactogenicity-type events and generally more common in the 18–55 years than the ≥56 years group in both booster cohorts (**Supplementary Table S6**).

**Figure 3.**
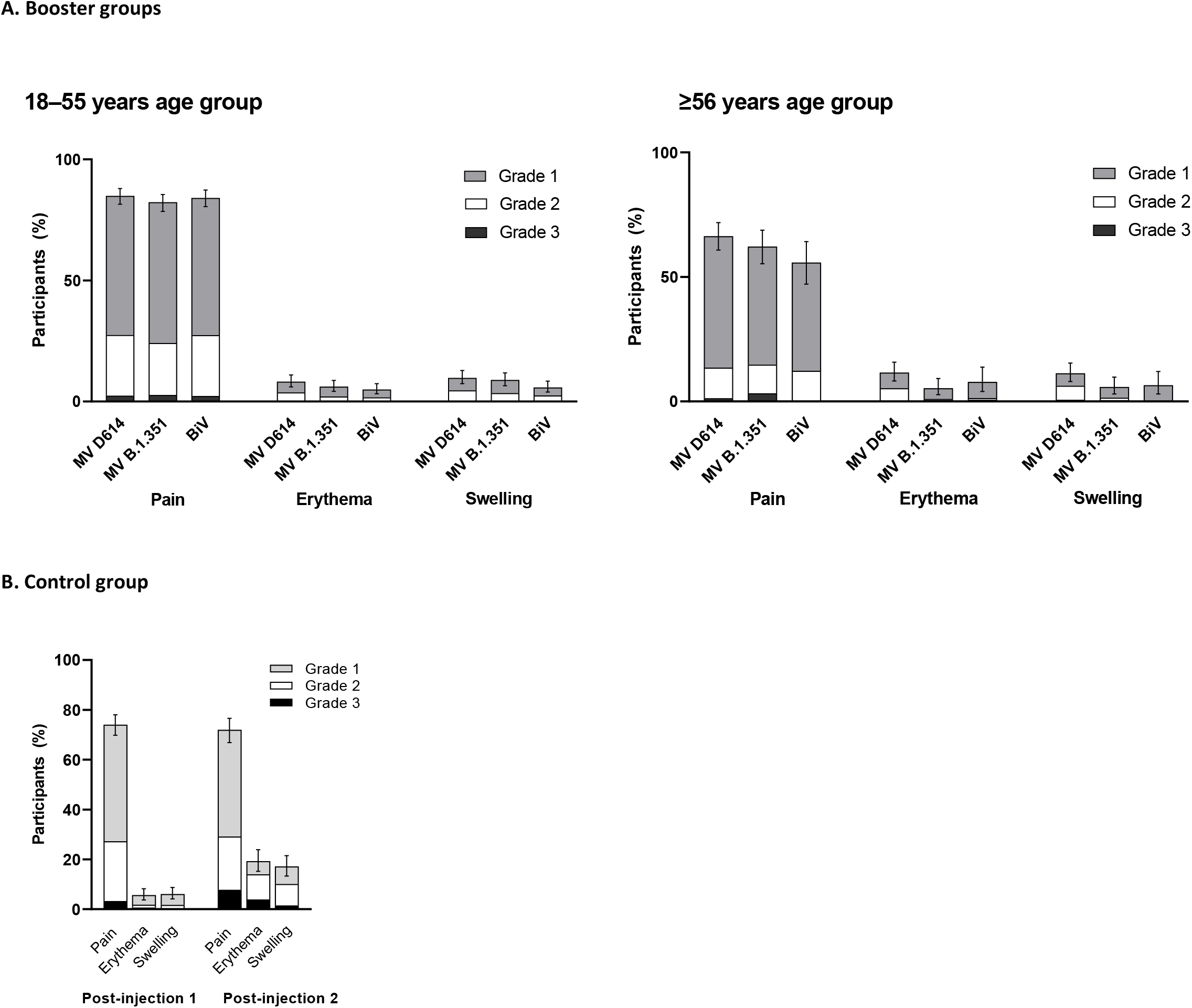
Solicited injection site reactions up to 7 days following vaccination in CoV2 preS dTM-AS03 booster groups and the control group, by age category - SafAS. The percentages of participants experiencing at least one of the specified reactions are shown. Error bars denote 95% CIs for the percentages after any injection, calculated using the Clopper-Pearson method. MV, monovalent; BiV, bivalent

**Figure 4.**
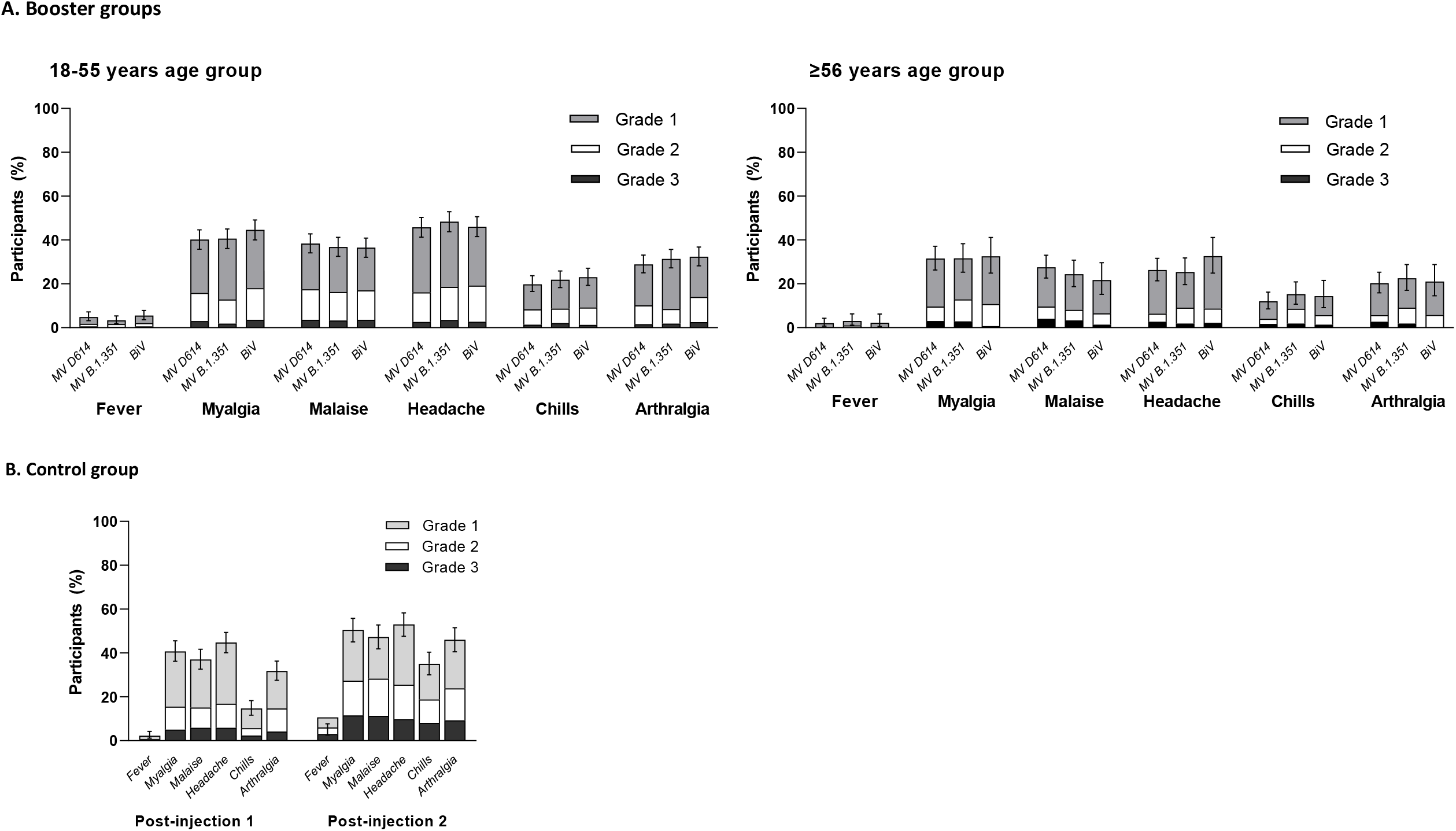
Solicited systemic reactions up to 7 days following vaccination in CoV2 preS dTM-AS03 booster groups and the control group, by age category – SafAS. The percentages of participants experiencing at least one of the specified reactions are shown. Error bars denote 95% CIs for the percentages after any injection, calculated using the Clopper-Pearson method. MV, monovalent; BiV, bivalent

One of 28 SAEs reported up to the analysis cut-off date was assessed as related to the study vaccine (serum sickness-like reaction in a participant primed with mRNA-1273 and boosted with bivalent D614+B.1.351), which resolved 11 days after onset following treatment (antihistamines and steroid cream). One participant aged ≥56 years, primed with CoV2 preS dTM-AS03 (D614) and boosted with monovalent B.1.351, reported an AESI (trigeminal neuralgia), assessed as unrelated to vaccination. Of 356 MAAEs reported by 264 booster recipients, eight (2 in the monovalent D614 group, 5 in the monovalent B.1.351 group and 1 in the bivalent D614+B.1.351 group) were assessed as related to the study vaccine. All MAAEs resolved. No deaths and no AEs leading to study discontinuation were reported. No cases of unintentional exposure during pregnancy were reported in any booster group.

The safety profile of the control group is summarized in **Supplementary Table S7**. Two participants reported grade 3 immediate unsolicited AEs. One had grade 3 rash, assessed as related and non-serious and resolved on the same day. Another had a grade 3 possible anaphylactic reaction immediately after the second dose (also reported as an AESI and a MAAE), which was assessed as related and resolved with treatment (intramuscular adrenaline and oral prednisone) on the same day. This case did not meet the Brighton Collaboration case definition for anaphylaxis.^23^ Solicited reactions were mostly mild to moderate and transient (**Figure 3**). Greater frequency and severity of solicited systemic reactions were reported post-dose 2 compared to post-dose 1, with grade 3 solicited systemic reactions reported for 56/456 (12.3%) participants post-dose 1 and 76/377 (20.2%) post-dose 2 (**Figure 4**). Unsolicited AEs were mostly of mild or moderate intensity and resolved within 7 days of onset. Eight participants reported eight SAEs in the control group, none assessed as related. One SAE (suicidal ideation) led to study discontinuation. A second AESI (grade 1 rheumatoid arthritis) was reported 73 days post-second dose, assessed as not related. Of 121 MAAEs reported by 84 participants over the analysis period, 15 (in 8 participants), were assessed as related; all resolved. No deaths were reported. One spontaneous abortion was reported in the control group, assessed as not related to the vaccine. No other exposure during pregnancy was reported in the control group.

## Discussion

The findings from this study show that CoV2 preS dTM-AS03 booster vaccines (monovalent D614, monovalent B.1.351 and bivalent D614+B.1.351) are well tolerated and elicit robust neutralizing antibody responses against variants including Omicron in younger and older adults regardless of the vaccine platform used for primary vaccination. Notably, greater neutralizing antibody responses against Omicron BA.2 were achieved after boosting with monovalent B.1.351 than the other formulations.

The cross-neutralization of Omicron subvariants following a Beta-variant containing vaccine observed in this study is consistent with initial findings from a phase 3 randomized trial, in which a booster dose of monovalent B.1.351 achieved higher neutralizing antibody titers against Beta, Delta and Omicron BA.1 variants than either a monovalent D614 booster or a third dose of BNT162b2, when administered to adults who previously received a primary vaccination series of BNT162b2.^24^ The cross-neutralization of Omicron subvariants with a Beta-variant containing vaccine has also been observed with the mRNA vaccine platform. A bivalent mRNA COVID-19 vaccine candidate encoding the ancestral SARS-CoV-2 and B.1.351 variant spike protein administered after a primary vaccination series with the original mRNA-1273 elicited robust cross-neutralizing antibodies against Omicron.^25^

This study benefits from a large global study population, inclusive of high-risk groups, thus closely representing the potential target populations. The inclusion of participants previously primed with different vaccine platforms (mRNA, adenovirus-vectored and adjuvant recombinant protein vaccines) including globally deployed vaccines, allowed a comprehensive analysis of prime-boosting options and make our findings applicable to a wide population of individuals. All three CoV2 preS dTM-AS03 booster formulations elicited robust neutralizing antibody responses irrespective of primary vaccination. While higher or similar responses have been previously reported for heterologous prime-booster regimens with mRNA or adenovirus-vectored vaccines compared to homologous prime-boost,^26-28^ the highest responses in the current study were observed following prime-boost with CoV2 preS dTM-AS03 vaccine formulations. These findings may be influenced by the variation in interval between primary and booster injections. Indeed, recent BNT162b2 booster vaccination data suggest that a longer interval (35 weeks or more) may increase vaccine effectiveness,^6^ and previous data have suggested that extended intervals between doses in a primary series of COVID-19 vaccination result in higher neutralizing antibody titers.^29^ Alternatively, the higher booster responses following CoV2 preS dTM-AS03 priming may result from differences in the priming of the immune system compared to the other priming vaccine platforms, or differences in the proportion of participants who had prior infection.

Although rare, higher frequencies than expected of myocarditis and pericarditis have been previously reported following receipt of COVID19 mRNA vaccines^30^ and with the adjuvanted, protein-based vaccine NVX-CoV2373^31,32^ and an increased risk of thrombosis with thrombocytopenia, following receipt of adenovirus-vectored vaccines.^33^ We did not identify any safety concerns in the current study. For all booster groups, reactogenicity tended to be transient, mild to moderate in severity, and similar (for solicited local reactions) or lower (for solicited systemic reactions) post-booster than post-primary vaccination series. The proportion of participants reporting a reactogenicity event after a CoV2 preS dTM-AS03 booster (any grade or grade 3) appears to be similar to or less than that observed after licensed COVID-19 boosters (BNT162b2, mRNA-1273, NVX-CoV2373).^34-36^

Some limitations should be noted. Due to a lack of data on previous infection, we cannot make meaningful direct comparisons across booster groups; also, we did not compare data for the ≥56 years age category to age-matched controls. The variability in the interval between last primary vaccine dose and booster may have affected the antibody responses to booster vaccination; similarly, the inclusion of immunocompromised participants in our study may have affected the immune responses observed. While our immunogenicity results are promising, it will be important to evaluate the durability of the responses observed in this study, beyond the 15 days post-booster period. Additionally, data on efficacy, as well as on cellular responses elicited by these vaccines, will be needed to understand their potential effectiveness against disease outcomes. Another limitation of our study is the lack of cross-neutralization data on BA.4/5 and more recent variants such as BQ.1.1 and BA.2.75. However, while at the time of writing, the dominant global VoC are Omicron subvariants BA.4 and BA.5,^37^ the nature of likely future SARS-CoV-2 variants remains unpredictable. Our data indicate that a vaccine containing a B.1.351-component can provide cross-neutralization against a heterologous emergent strain and challenges the current variant-chasing vaccine paradigm.

In summary, B.1.351-containing CoV2 preS dTM-AS03 vaccines given as a booster following primary vaccination with globally deployed vaccines elicited strong cross-neutralizing antibody responses against SARS-CoV-2 variants, including against Omicron variants, and had acceptable safety profiles. These findings support the deployment of monovalent B.1.351 as a universal booster against COVID-19.

## Data Availability

Qualified researchers can request access to patient-level data and related study documents, including the clinical study report, study protocol with any amendments, blank case report forms, statistical analysis plan, and dataset specifications. Patient-level data will be anonymised and study documents will be redacted to protect the privacy of trial participants. Further details on Sanofi's data sharing criteria, eligible studies, and process for requesting access can be found at https://vivli.org/.

## Author Contributions

GdB, MAC, FTDS, BF, MHG, OH, MK, NLM, RMC, NR, LS, SG, SSa and SSr contributed to the concept or design of the study and data analysis and interpretation; MIB and RM contributed to the conception or design of the study and data acquisition; MSR, HA, SA, DB, AB, RC, SD, DD, AF, RF, JG, PG, DH, OL, JAH, FMT, JP, DMRM, CR, LDS and AW contributed to data acquisition; AP and JW were involved in the analysis and interpretation of the data. All authors were involved in drafting or critically revising the manuscript, and all authors approved the final version and are accountable for the accuracy and integrity of the manuscript. All authors had full access to all the data and accept responsibility to submit for publication.

## Declaration of Interests

GdB, JW, AP, MSR, HA, MIB, RC, RF, BF, JG, M-HG, SG, OH, RM, JP, NR, RMC and SSa are Sanofi employees; MIB, RMC hold stock options. SSa was a Sanofi employee at the time of study conduct. SSr, RMC, GdB, CAD and SSa, are inventors on a pending patent application filed by Sanofi and GSK for the development of the CoV-2 dTM vaccine. MAC, LS and MK are employed by GSK and hold restricted shares in the GSK group of companies. FTDS was employed by GSK, and held restricted shares in the GSK group of companies, at the time of the study. DMRM declares that her institution received funding from Sanofi. FM-T has received honoraria from GSK group of companies, Pfizer Inc, Sanofi, MSD, Seqirus, Biofabri, and Janssen for taking part in advisory boards and expert meetings and for acting as a speaker in congresses outside the scope of the submitted work. FM-T has also acted as principal investigator in randomized controlled trials of the above-mentioned companies as well as Ablynx, Gilead, Regeneron, Roche, Abbott, Novavax, and MedImmune, with honoraria paid to his institution. AF receives research funding, paid to his employers, from Sanofi both for work related to this study and other unrelated vaccine trials and from GSK for other unrelated studies. He receives research funding from other vaccine manufacturers relating to trials and studies and undertakes paid consultancy related to a number of developmental antimicrobial drugs and vaccines.

## Acknowledgements

The authors thank all participants, investigators, and study site personnel who took part in this study. The authors acknowledge Juliette Gray of inScience Communications, Springer Healthcare, London, UK, for providing editorial assistance with the preparation of this manuscript, funded by Sanofi. The authors also thank Hanson Geevarghese for providing manuscript coordination on behalf of Sanofi. This work was done in collaboration with GSK, who provided access to, and use of, the AS03 Adjuvant System.

## Role of the funding source

Funding was provided by Sanofi and by federal funds from the Biomedical Advanced Research and Development Authority, part of the office of the Administration for Strategic Preparedness and Response at the U.S. Department of Health and Human Services under Contract # HHSO100201600005I, and in collaboration with the U.S. Department of Defense Joint Program Executive Office for Chemical, Biological, Radiological and Nuclear Defense under Contract # W15QKN-16-9-1002. The funders were involved in the study design, data collection, data analysis, data interpretation, writing of the report, and the decision to submit the paper for publication. GSK provided access to, and use of, the AS03 Adjuvant System.

## Notes

### Clinical Trial

NCT04762680

### Author Declarations

The study was undertaken in compliance with the International Conference on Harmonization guidelines for Good Clinical Practice and the principles of the Declaration of Helsinki. The protocol and amendments were approved by Independent Ethics Committees (country ethics committees for Honduras [Comite Etica Independiente Zugueme], France [Comite De Protection Des Personnes Ile De France III Hopital Tarnier-Cochin, France], Spain [Comite De Eticade Investigacion Con Medicamentos Parc Tauli, Spain] and the UK [HRA and Health and Care Research Wales, UK], a central ethics committee for the USA [Advarra] and local ethics committees for the USA [WCG IRB, Columbia Research Human Research Protection Office, Langone Health Office of Science and Research Institutional Review Board and the Yale Human Research Protection Program] and for Australia [Bellberry, Sydney Children's Hospitals Network Human Research Ethics Committee and Sydney Children's Hospital Network (SCHN) Research Governance]) as per local regulations. Written informed consent was obtained from all participants before any study procedures were performed.

