## Supplementary Material for "Safety and immunogenicity of a variant-adapted SARS-CoV-2 recombinant protein vaccine with AS03 adjuvant as a booster in adults primed with authorized vaccines"

Guy de Bruyn, Joyce Wang, Annie Purvis, Martin Sanchez Ruiz, Haritha Adhikarla, Saad Alvi, Matthew I Bonaparte, Daniel Brune, Agustin Bueso, Richard M Canter, Maria Angeles Ceregido, Sachin Deshmukh, David Diemert, Adam Finn, Remi Forrat, Bo Fu, Julie Gallais, Paul Griffin, Marie-Helene Grillet, Owen Haney, Jeffrey A Henderson, Marguerite Koutsoukos, Odile Launay, Federico Martinon Torres, Roger Masotti, Nelson L Michael, Juliana Park, Doris M Rivera M, Natalya Romanyak, Chris Rook, Lode Schuerman, Lawrence D Sher, Fernanda Tavares-Da-Silva, Ashley Whittington, Roman M Chicz, Sanjay Gurunathan, Stephen Savarino, Saranya Sridhar; Allaw, Mohammed; Babin, Valérie; Babyak, Jennifer; Ben-Ghezala, Ines; Breuer, Thomas; Breymeier, Corinne; Conrad, Anne; Holmqvist, Ciarrah; Costa-Araujo, Cristiana; Coux, Florence; Dellanno, Christine; Dussol, Bertrand; Essink, Brandon; Garrido, Jesús; Girodet, Pierre-Olivier; Gonzalez, Claudia; Grosbois, Marie-Ange; Hammond, Justin; He, Chelsea; Homlqvist, Ciarrah; Hudzina, Kathy; Hutchens, Mark; Jackson Booth, Peta-Gay; Joaquin, Arnel; Kandasamy, Rama; Kasztejna, Jennifer; Keefer, Michael; Kimmel, Murray; Kresge, Matthew; Laine, Fabrice; Lefebvre, Maeva; Lopez, Denise; Lourdes, Malaborbor Perpetua; Maakaroun-Vermesse, Zoha; Malishchak, Caitlin; Menard, Lisa; Mendoza, Sandra; Moore, Patrick; Mulamalla, Mounika; Mulholland, Patrick; Nicolas, Jean-Francois; Ogbuagu, Onyema; Ortiz, Juan; Perroud, Ana Paula; Peyton, Gina; Purvis, Ya-Fen; Raabe, Vanessa; Rivas, Enrique; Rouphael, Nadine; Roy, Beatrice; Sagot, Lola; Sater, Nessryne; Schwartz, Howard; Severance, Randall; Shi, Jiayuan; Sobieszczyk, Magdalena; Stevens, Charlene; Thuy, Tran, Phuong; Toma, Ramy; Tong, Tina; Tourneux, Sophie; Treanor, John ; Turet, Núria; Froget, Rachel; Walsh, Stephen; White, Judith; del Campo Perez, Victor; Perez Breva, Lina; Rojo Conejo, Pablo; Ruiz Antoraz, Maria Belen; Chin, Toong; Fribbens, Charlotte; Phillipson, Adrian; Kaminski, Rachel; Emmett, Stevan; Hebert, Corey; Birch, Thomas; Roberson, Russell; Zacher, Jeffrey; Gelu-Maury, Sophie; Loryne, Loron; Davis, Yvonne

### 2 Study site investigators

*Australia*: Rama Kandasamy;

*France*: Ines Ben-Ghezala, Anne Conrad, Bertrand Dussol, Pierre-Olivier Girodet, Fabrice Laine, Maeva Lefebvre, Zoha Maakaroun-Vermesse, Jean-Francois Nicolas, Rachel Froget;

*Spain*: Victor del Campo Perez, Pablo Rojo Conejo, Maria Belen Ruiz Antoran

*UK*: Justin Hammond, Patrick Moore, Toong Chin, Charlotte Fribbens, Adrian Phillipson, Rachel Kaminski, Stevan Emmett;

*USA*: Mohammed Allaw, Brandon Essink, Mark Hutchens, Peta-Gay Jackson Booth, Arnel Joaquin, Michael Keefer, Murray Kimmel, Onyema Ogbuagu, Vanessa Raabe, Nadine Rouphael, Howard Schwartz, Randall Severance, Magdalena E Sobieszczyk, Ramy Toma, Stephen Walsh, Judith White, Corey Hebert, Thomas Birch, Russell Roberson, Jeffrey Zacher.

### 3 Supplementary Methods

#### 3.1 Participant inclusion/exclusion criteria

Adults, aged 18 years and older, were eligible. Exclusion criteria included women who were pregnant or lactating, or, for those of childbearing potential, not using an effective method of contraception or abstinence from at least 4 weeks prior to the first vaccination until at least 12 weeks after the last vaccination; known systemic hypersensitivity to any of the vaccine components, or history of a life-threatening reaction to a vaccine containing any of the same substances; dementia or any other cognitive condition at a stage that could interfere with following the trial procedures based on Investigator or designee's judgment; self-reported thrombocytopenia, contraindicating intramuscular (IM) vaccination based on Investigator or designee's judgment; bleeding disorder, or receipt of anticoagulants in the past 21 days preceding inclusion, contraindicating IM vaccination based on Investigator or designee's judgment; unstable acute or chronic illness that in the opinion of the Investigator or designee poses additional risk as a result of participation or that could interfere with the trial procedures; receipt of solid-organ or bone marrow transplants in the past 180 days; receipt of anti-cancer chemotherapy in the last 90 days; moderate or severe acute illness/infection (according to investigator judgment) on the day of vaccination or febrile illness (temperature ≥38.0°C [≥100.4°F]); receipt of any vaccine in the 30 days preceding or on the day of the first trial vaccination or planned receipt of any vaccine in the 30 days following the second trial vaccination except for influenza vaccination, which could be received at any time in relation to trial intervention; participation at the time of trial enrollment (or in the 30 days preceding the first trial vaccination) or planned participation during the present trial period in another clinical trial investigating a vaccine, drug, medical device, or medical procedure. Additionally, in the control group, participants were not included if they had prior administration of a coronavirus vaccine (SARS-CoV-2, SARS-CoV, Middle East Respiratory Syndrome [MERS-CoV]).

To allow evaluation of vaccine performance in high-risk groups, individuals with other high-risk medical conditions (including those considered to potentially increase the risk for severe COVID-19 illness) or those who were immunocompromised for other reasons, were not excluded from the study.

#### 3.2 Study vaccine and vaccine administration

The recombinant protein antigen CoV2 preS dTM and the AS03 Adjuvant System (GSK Vaccines, Rixensart, Belgium) have been described previously.^23,42^ The CoV2 preS dTM vaccine and AS03 adjuvant formulations were presented in two separate vials: a multi-dose vial containing AS03 (sufficient for 10 doses) and a single-dose vial containing CoV2 preS dTM antigen. An equal volume of the adjuvant emulsion was added to the vial containing the antigen and mixed prior to injection. Vaccinations (0.5 mL per dose) were administered on study day 1 (booster dose) or on days 1 and 22 (primary doses) by qualified and trained personnel by intramuscular injection into the deltoid region of the upper arm.

The CoV2 preS dTM-AS03 booster formulations evaluated in this study each contained a total of 5 µg of CoV2 preS dTM antigen, as follows: monovalent D614 contained 5 µg of the D614 antigen; monovalent B.1.351 contained 5 µg of the B.1.351 antigen; and bivalent D614+B.1.351 contained 2.5 µg of D614 plus 2.5 µg of B.1.351.

#### 3.3 Measurement of SARS-CoV-2 neutralising antibody titers using a pseudovirus neutralization assay (Monogram)

SARS-CoV-2 neutralizing antibody (NAb) titers against the D614G strain or B.1.351 or Omicron BA.1 and BA.2 subvariants were measured using a pseudovirus neutralization (PsVN) assay performed at Monogram Biosciences LabCorp (South San Francisco, CA, USA). Pseudoviruses were prepared by co-transfecting HEK293 producer cells with an HIV-1 genomic vector and a SARS CoV-2 envelope expression vector. NAb activity was measured by assessing the inhibition of luciferase activity in HEK293 target cells expressing the ACE2 receptor following pre-incubation of the pseudovirions with serial dilutions of the serum specimen. The expression of luciferase activity in target cells is inhibited in the presence of anti- SARS-CoV-2 NAb. Data were displayed by plotting the percent inhibition of luciferase activity versus the log_10_ reciprocal of the serum/plasma dilution and NAb titers were reported as the reciprocal of the serum dilution conferring 50% inhibition (ID_50_) of pseudovirus infection. To ensure that the measured NAb activity was SARS-CoV-2 specific, each test specimen was also assessed using a non-specific pseudovirus (specificity control) that expresses a non-reactive envelope protein of one or more unrelated viruses.

#### 3.5 Primary and secondary objectives evaluated for each booster group

| **Co-primary immunogenicity objectives-** | |
| --- | --- |
| *Cohort 1* | |
|  | **1.** To demonstrate in adults 18-55 years of age and previously vaccinated with the Pfizer/BioNTech mRNA COVID-19 vaccine that a booster dose of CoV2 preS dTM-AS03 (D614) vaccine induces an immune response that is non-inferior to the response induced by a two ‑dose priming series with the CoV2 preS dTM-AS03 (D614) vaccine in naïve, previously unvaccinated individuals |
|  | **2.** To demonstrate in adults 18-55 years of age and previously vaccinated with the Pfizer/BioNTech mRNA COVID-19 vaccine that a booster dose of CoV2 preS dTM-AS03 (D614) vaccine induces an immune response that is superior to that observed immediately before booster |
| *Cohort 2* | |
|  | **1.** To demonstrate in adults 18-55 years of age and previously vaccinated with the Pfizer/BioNTech mRNA COVID-19 vaccine that a booster dose of CoV2 preS dTM-AS03 (B.1.351) or CoV2 preS dTM-AS03 (D614 + B.1.351) induces an immune response that is non-inferior to the response induced by a two-dose priming series with the CoV2 preS dTM-AS03 (D614) vaccine in naïve, previously unvaccinated individuals |
|  | **2.** To demonstrate in adults 18-55 years of age and previously vaccinated with the Pfizer/BioNTech mRNA COVID-19 vaccine that a booster dose of CoV2 preS dTM-AS03 (B.1.351) or CoV2 preS dTM-AS03 (D614 + B.1.351) induces an immune response that is superior to that observed immediately before booster |
| **Secondary objectives for cohort 1** | |
| *Conditional on meeting the primary objectives for cohort 1* | |
|  | **1.** To demonstrate, in adults 18-55 years of age, previously vaccinated with the CoV2 preS dTM-AS03 (D614) vaccine, that a booster dose of CoV2 preS dTM-AS03 (D614) vaccine induces an immune response that is non-inferior to the response induced by a two-dose priming series with CoV2 preS dTM-AS03 (D614) vaccine in naïve, previously unvaccinated, individuals |
|  | **2.** To demonstrate, in adults 18-55 years of age, previously vaccinated with CoV2 preS dTM-AS03 (D614) vaccine, that a booster dose of CoV2 preS dTM-AS03 (D614) induces an immune response that is superior to that observed immediately before booster |
| *Conditional on meeting the conditional secondary objectives (1) and (2) for cohort 1* | |
|  | **3.** To demonstrate in adults 18-55 years of age and previously vaccinated with an mRNA COVID-19 vaccine, that a booster dose of CoV2 preS dTM-AS03 (D614) vaccine induces an immune response that is non-inferior to the response induced by a two-dose priming series with CoV2 preS dTM-AS03 (D614) vaccine in naïve, previously unvaccinated individual |
|  | **4.** To demonstrate in adults 18-55 years of age and previously vaccinated with an mRNA COVID-19 vaccine, that a booster dose of CoV2 preS dTM-AS03 (D614) vaccine induces an immune response that is superior to that observed immediately before booster. |
| *Conditional on meeting the conditional secondary objectives (3) and (4) for cohort 1* | |
|  | **5.** To demonstrate, in adults 18-55 years of age previously vaccinated with an adenovirus-vectored COVID-19 vaccine, that a booster dose of CoV2 preS dTM-AS03 (D614) vaccine induces an immune response that is non-inferior to the response induced by a two-dose priming series with CoV2 preS dTM-AS03 (D614) vaccine in naïve, previously unvaccinated, individuals |
|  | **6.** To demonstrate, in adults 18-55 years of age previously vaccinated with adenovirus-vectored COVID-19 vaccines, that a booster dose of CoV2 preS dTM-AS03 (D614) induces an immune response that is superior to that observed immediately before booster |
| *Other secondary objectives for cohort 1* | |
| To describe, in adults 18-55 years of age previously vaccinated with the Moderna, Oxford/Astra-Zeneca, or Janssen COVID-19 mRNA vaccine, the immune response to a booster dose of CoV2 preS dTM-AS03 (D614). | |
| To compare the neutralizing antibody profile to the B.1.351 variant at study day (D)1 and D15 in the booster groups with the neutralizing antibody profiles to the D614G and B.1.351 strains at D36 in the Comparator group. | |
| **Secondary objectives for cohort 2** | |
| *Conditional on meeting the primary objectives for cohort 2* | |
|  | **1.** To demonstrate in adults 18-55 years of age and previously vaccinated with an mRNA COVID-19 vaccine, that a booster dose of CoV2 preS dTM-AS03 (B.1.351) or CoV2 preS dTM-AS03 (D614 + B.1.351) vaccine induces an immune response that is non-inferior to the response induced by a two-dose priming series with CoV2 preS dTM-AS03 (D614) vaccine in naïve, previously unvaccinated individuals.  **2.** To demonstrate in adults 18-55 years of age and previously vaccinated with an mRNA COVID-19 vaccine, that a booster dose of CoV2 preS dTM-AS03 (B.1.351) or CoV2 preS dTM-AS03 (D614 + B.1.351) induces an immune response that is superior to that observed immediately before booster |
| *Conditional on meeting the the co-secondary objectives for cohort 2* | |
|  | **3.** To demonstrate in adults 18-55 years of age and previously vaccinated with the Pfizer/BioNTech mRNA COVID-19 vaccine, that a booster dose of CoV2 preS dTM-AS03 (B.1.351) or CoV2 preS dTM-AS03 (D614 + B.1.351) induces an immune response against the B.1.351 variant at D15 that is superior to that against the B.1.351 variant at D36 in the Comparator Group |
| *Other secondary objectives for cohort 1* | |
|  | To describe, in adults 18-55 years of age and previously vaccinated with an adenovirus-vectored COVID-19 vaccine, the immune response to a booster dose of CoV2 preS dTM-AS03 (B.1.351) or CoV2 preS dTM-AS03 (D614 + B.1.351) vaccine compared to the response induced by a two dose priming series with CoV2 preS dTM-AS03 (D614) vaccine in naïve, previously unvaccinated individuals. |
|  | To describe, in adults 18-55 years of age and previously vaccinated with an adenovirus-vectored COVID-19 vaccine, the immune response to a booster dose of CoV2 preS dTM-AS03 (B.1.351) or CoV2 preS dTM-AS03 (D614 + B.1.351) compared to that observed immediately before booster. |
|  | To describe, in adults 18-55 years of age previously vaccinated with the Moderna, Oxford/Astra-Zeneca or Janssen COVID-19 mRNA vaccine, the immune response to a booster dose of CoV2 preS dTM-AS03 (B.1.351) or CoV2 preS dTM-AS03 (D614 + B.1.351). |
|  | To compare the neutralizing antibody profile to the B.1.351 variant at D15 in each study Intervention Group and D36 in the Comparator Group, overall, by priming platform, and by priming vaccine. |
|  | To compare the neutralizing antibody profile against the D614G strain at D15 following a booster dose of CoV2 preS dTM-AS03 (D614 + B.1.351) and at D36 in the Comparator Group. |

The full list of study objectives can be found in the study protocol.

#### 3.6 Statistics

Log-transformed individual titers (log_10_(data)), assumed to be normally distributed, were used to calculate geometric mean titers (GMTs) and GMT ratios. Two-sided 98.3% CIs and 95% CIs were calculated based on the Student t-distribution of log_10_ titers for within-group comparisons and on Welch’s t-interval for between-group comparisons (booster group versus control). Statistical inference was based on the use of two-sided 98.3% CIs. Non-inferiority in terms of neutralizing antibody titers was concluded if the lower bound of the 98.3% CI for the between-group (booster versus control) GMT ratio was >0.67. Superiority in terms of neutralizing antibody titers was concluded if the lower limit of the 2-sided 98.3% CI of the fold-rise (post-booster versus pre-booster) was >2. Non-inferiority in terms of seroresponse rates was concluded if the lower limit of the 2-sided 98.3% CI of the difference of seroresponse between groups (booster – control) was ≥−10%. To correct for multiple co-primary endpoints, Bonferroni adjustment was used to correct for Type I error at 0.025 (one-sided), with each comparison of the booster group to the control group performed at a one-sided significance level of 0.00833.

Planned sample sizes with ≥90% power overall for the 18–55 years group were: 215 BNT162b2-primed, 270 CoV2 preS dTM-AS03 (D614)-primed participants and 75 mRNA-1273-primed participants in the monovalent D614 booster group; 515 BNT162b2-primed and 75 mRNA-1273-primed participants in both the monovalent B.1.351 and bivalent D614+B.1.351 booster groups; and 515 SARS-CoV-2 naïve participants in the control group. The assumed attrition rates were 3% in the booster cohorts and 10% in the control group. For descriptive analyses, 75 participants 18–55 years of age and 25 participants ≥56 years of age for each remaining priming vaccine (other than those described above), were planned to be enrolled into each booster group. To enable extrapolation, approximately 50 BNT162b2-primed participants ≥56 years of age were planned to be enrolled into each booster group. In the older adult stratum, to extend the characterization of the product, enrolment was allowed to continued and was not capped in cases where the planned numbers were achieved. Among CoV2 PreS dTM-AS03-primed participants, approximately 300 in the 18-55 years age group and 150 in the ≥56 years age group were assumed to transition from the original phase 2 study into the monovalent D614 and monovalent B.1.351 groups.

#### 3.7 Determination of SARS-CoV-2 naïve status for immunogenicity assessment in the control group

In the control group, SARS-CoV-2 naïve status was determined on serological samples using ELECSYS electrochemiluminescence immunoassays for anti-SARS-CoV-2 anti-S (S-ELECSYS; Elecsys Anti-SARS-CoV-2 S, Roche, Indianapolis, IN, USA EUA202698) on study day (D)1 and on nasopharyngeal swabs using nucleic acid amplification tests (NAAT; Abbott RealTime SARS-CoV-2 assay, EUA200023) on D1 and D22, following the manufacturer’s instructions. Participants were categorized as naïve if both tests showed negative results on both D1 and D2.

### 4 Supplementary Results

#### 4.1 Safety after primary vaccination with CoV2 PreS dTM-AS03 (10 ug) in the control group.

See **Table S7** for a safety overview after any injection in the control group. Two participants reported grade 3 immediate unsolicited AEs. One had grade 3 rash, assessed as related and non-serious and resolved on the same day. Another had a grade 3 possible anaphylactic reaction immediately after the second dose, which was assessed as related and resolved with treatment (intramuscular adrenaline and oral prednisone) on the same day (also reported as an AESI). This case did not meet the Brighton Collaboration case definition for anaphylaxis.^1^

### 5 Supplementary Tables and Figures

**Table S1. High-risk medical conditions of study participants (SafAS)**

|  | **CoV2 preS dTM-AS03 booster formulation** | | | **Control** |
| --- | --- | --- | --- | --- |
|  | **Monovalent D614**  **N=803** | **Monovalent B.1.351 N=705** | **Bivalent D614+B.1.351 N=621** | **(two CoV2 preS dTM-AS03 D614 primary vaccine doses)  N=473** |
| **Participants with the high-risk medical condition, n (%)** | **n (%)** | **n (%)** | **n (%)** | **n (%)** |
| Obesity (body mass index of 30 or higher) | 234 (29.1) | 257 (36.5) | 216 (34.8) | 169 (35.7) |
| Smoking | 66 (8.2) | 132 (18.7) | 125 (20.1) | 145 (30.7) |
| Cancer | 36 (4.5) | 20 (2.8) | 12 (1.9) | 9 (1.9) |
| Chronic Kidney Disease | 4 (0.5) | 1 (0.1) | 3 (0.5) | 0 |
| Immunocompromised State from Solid Organ Transplantation | 1 (0.1) | 0 | 0 | 0 |
| Immunocompromised State from Other Causes | 21 (2.6) | 12 (1.7) | 10 (1.6) | 5 (1.1) |
| Hepatic Disease | 9 (1.1) | 5 (0.7) | 4 (0.6) | 2 (0.4) |
| Neurologic Conditions | 11 (1.4) | 15 (2.1) | 16 (2.6) | 8 (1.7) |
| Type 1 Diabetes Mellitus | 3 (0.4) | 9 (1.3) | 5 (0.8) | 1 (0.2) |
| Type 2 Diabetes Mellitus | 58 (7.2) | 69 (9.8) | 31 (5.0) | 15 (3.2) |
| Chronic Obstructive Pulmonary Disease | 10 (1.2) | 8 (1.1) | 7 (1.1) | 3 (0.6) |
| Cystic Fibrosis | 2 (0.2) | 0 | 0 | 0 |
| Pulmonary Fibrosis | 0 | 1 (0.1) | 1 (0.2) | 1 (0.2) |
| Moderate to Severe Asthma | 23 (2.9) | 30 (4.3) | 28 (4.5) | 10 (2.1) |
| Sickle Cell Disease | 0 | 0 | 0 | 0 |
| Thalassemia | 0 | 1 (0.1) | 2 (0.3) | 1 (0.2) |
| Heart Conditions such as Heart Failure | 16 (2) | 11 (1.6) | 11 (1.8) | 6 (1.3) |
| Coronary Artery Disease or Cardiomyopathies | 18 (2.2) | 16 (2.3) | 7 (1.1) | 1 (0.2) |
| Hypertension / High Blood Pressure | 172 (21.4) | 144 (20.4) | 100 (16.1) | 28 (5.9) |
| Cerebrovascular Disease | 10 (1.2) | 5 (0.7) | 2 (0.3) | 0 |

High-risk conditions are those considered to be associated with an increased risk of severe COVID-19, as detailed at https://www.cdc.gov/coronavirus/2019-ncov/science/science-briefs/underlying-evidence-table.html; ^†^Obesity: body mass index of 30 kg/m^2^ or higher

**Table S2.** Pseudovirus neutralising antibody responses against D614G 14 days after a monovalent D614 booster dose - PPAS

**S2.A. By priming vaccine and age group**

|  | **BNT162b2-primed** **(N=302)** | | **mRNA-1273-primed (N=102)** | | **ChAdOx1nCoV-19-primed (N=119)** | | **Ad26.CoV2.S-primed (N=92)** | | **CoV2 preS dTM-AS03 (D614)-primed (N=119)** | |
| --- | --- | --- | --- | --- | --- | --- | --- | --- | --- | --- |
|  | **M** | **GMT, GMTR or %** | **M** | **GMT, GMTR or %** | **M** | **GMT, GMTR or %** | **M** | **GMT, GMTR or %** | **M** | **GMT, GMTR or %** |
|  |  | **(95% CI)** |  | **(95% CI)** |  | **(95% CI)** |  | **(95% CI)** |  | **(95% CI)** |
| **18–55 years** |  |  |  |  |  |  |  |  |  |  |
| GMT D1 | 189 | 339 (285 ; 402) | 66 | 808 (634 ; 1030) | 58 | 249 (165 ; 376) | 50 | 224 (140 ; 357) | 45 | 510 (274 ; 950) |
| GMT D15 | 202 | 7894 (6993 ; 8911) | 67 | 6958 (5562 ; 8705) | 68 | 6663 (5366 ; 8274) | 63 | 7204 (5587 ; 9290) | 52 | 26584 (21800 ; 32418) |
| GMTR D15/D1 | 189 | 23.4 (19.4 ; 28.2) | 66 | 8.67 (6.44 ; 11.7) | 58 | 27.3 (18.2 ; 40.9) | 50 | 34.0 (20.6 ; 56.1) | 45 | 51.6 (26.1 ; 102) |
| ≥4-fold rise* | 176/189 | 93.1 (88.5 ; 96.3) | 47/66 | 71.2 (58.7 ; 81.7) | 50/58 | 86.2 (74.6 ; 93.9) | 45/50 | 90.0 (78.2; 96.7) | 36/45 | 80.0 (65.4 ; 90.4) |
| **≥ 56 years** |  |  |  |  |  |  |  |  |  |  |
| GMT D1 | 89 | 230 (168 ; 316) | 32 | 473 (290 ; 774) | 43 | 222 (128 ; 383) | 25 | 259 (124 ; 541) | 53 | 157 (87.8 ; 280) |
| GMT D15 | 99 | 5392 (4427 ; 6567) | 34 | 5294 (3552 ; 7891) | 50 | 5744 (4444 ; 7423) | 28 | 6188 (4239 ; 9035) | 66 | 19918 (15622 ; 25395) |
| GMTR D15/D1 | 89 | 22.9 (16.0 ; 33.0) | 32 | 10.7 (6.28 ; 18.4) | 43 | 25.8 (15.4 ; 43.0) | 25 | 25.3 (11.9 ; 53.6) | 53 | 127 (67.7 ; 237) |
| ≥4-fold rise* | 76/89 | 85.4 (76.3 ; 92.0) | 29/32 | 71.9 (53.3; 86.3) | 37/43 | 86.0 (72.1 ; 94.7) | 21/25 | 84.0 (63.9 ; 95.5) | 48/53 | 90.6 (79.3 ; 96.9) |

**S2.B. By priming vaccine platform and age group**

|  | **mRNA-priming platform** | | | | **Adenovirus-vectored priming platform** | | | |
| --- | --- | --- | --- | --- | --- | --- | --- | --- |
|  | **18–55 years  N=269** | | **≥ 56 years N=135** | | **18–55 years N=132** | | **≥ 56 years N=79** | |
|  | **M** | **GMT, GMTR or %** | **M** | **GMT, GMTR or %** | **M** | **GMT, GMTR or %** | **M** | **GMT, GMTR or %** |
|  |  | **(95% CI)** |  | **(95% CI)** |  | **(95% CI)** |  | **(95%)** |
| GMT D1 | 255 | 424 (365 ; 492) | 121 | 279 (213 ; 365) | 108 | 237 (175 ; 321) | 68 | 235 (153 ; 360) |
| GMT D15 | 269 | 7650 (6879 ; 8507) | 133 | 5367 (4501 ; 6400) | 131 | 6918 (5872 ; 8151) | 78 | 5900 (4790 ; 7266) |
| GMTR D15/D1 | 255 | 18.1 (15.3 ; 21.4) | 121 | 18.8 (13.8 ; 25.4) | 108 | 30.2 (22.0 ; 41.3) | 68 | 25.6 (16.9 ; 38.7) |
| ≥4-fold rise* | 223/255 | 87.5 (82.7 ; 91.3) | 99/121 | 81.8 (73.8 ; 88.2) | 95/108 | 88.0 (80.3 ; 93.4) | 58/68 | 85.3 (74.6 ; 92.7) |

*Seroresponse (defined as ≥4-fold rise in neutralizing titer, post-/pre-vaccination)

CI, confidence interval; D, study day; GMT, geometric mean titer; GMTR (geometric mean titer ratio): geometric mean of individual titer ratios (post-vaccination/pre-vaccination); M: number of participants with available data for the relevant endpoint; N: number of participants in PPAS

2-sided 95% CI for GMT is based on the Student t-distribution of logarithmic transformation of the individual titers. 2-sided exact 95% CI for percentage of participants with 4-fold rise is based on the Clopper-Pearson method.

**Table S3. Pseudovirus neutralising antibody responses after one or two doses of CoV2 preS dTM-AS03 primary vaccination in the control group – PPAS naïve at D1+D22, by age group**

|  | **GMT** | | **GMTR** | | **≥4-fold rise*** | |
| --- | --- | --- | --- | --- | --- | --- |
|  | **M** | **1/dilution (95% CI)** | **M** | **Ratio (95% CI)** | **n/M** | **% (95% CI)** |
| **All ages** |  |  |  |  |  |  |
| *D614G PsVN* |  |  |  |  |  |  |
| D1 | 336 | 20.3 (19.7 ; 21.0) |  | - | - | - |
| D22 | 286 | 52.4 (44.0 ; 62.5) | 286 | 2.57 (2.16 ; 3.06) | 88/286 | 30.8 (25.5 ; 36.5) |
| D36 | 307 | 3611 (3086 ; 4224) | 306 | 178 (151; 211) | 303/306 | 99.0 (97.2 ; 99.8) |
| *B.1.351 PsVN* |  |  |  |  |  |  |
| D1 | 335 | 20.3 (19.7 ; 20.8) |  | - | - | - |
| D22 | 332 | 29.7 (26.0 ; 34.0) | 331 | 1.47 (1.29 ; 1.67) | 33/331 | 10.0 (7.0 ; 13.7) |
| D36 | 296 | 413 (346 ; 493) | 295 | 20.3 (16.9 ; 24.3) | 255/295 | 86.4 (82.0; 90.1) |
| **18–55 years** |  |  |  |  |  |  |
| *D614G PsVN* |  |  |  |  |  |  |
| D1 | 330 | 20.3 (19.7 ; 21.0) |  | - | - | - |
| D22 | 281 | 53.3 (44.6 ; 63.8) | 281 | 2.62 (2.19 ; 3.12) | 88/281 | 31.3 (25.9 ; 37.1) |
| D36 | 302 | 3658 (3123 ; 4286) | 301 | 181 (152 ; 214) | 298/301 | 99.0 (97.1 ; 99.8) |
| *B.1.351 PsVN* |  |  |  |  |  |  |
| D1 | 329 | 20.3 (19.7 ; 20.8) |  | - |  | - |
| D22 | 326 | 29.4 (25.7 ; 33.5) | 325 | 1.45 (1.28 ; 1.65) | 32/325 | 9.8 (6.8 ; 13.6) |
| D36 | 291 | 413 (346 ; 493) | 290 | 20.3 (16.9 ; 24.3) | 251/290 | 86.6 (82.1 ; 90.3) |
| **≥56 years** |  |  |  |  |  |  |
| *D614G PsVN* |  |  |  |  |  |  |
| D1 | 6 | 20.0 (NC ; NC) |  | - | - | - |
| D22 | 5 | 20.0 (NC ; NC) | 5 | 1.00 (NC ; NC) | 0/5 | 0 (0 ; 52.2) |
| D36 | 5 | 1640 (NC ; NC) | 5 | 82.0 (NC ; NC) | 5/5 | 100 (47.8 ; 100) |
| *B.1.351 PsVN* |  |  |  |  |  |  |
| D1 | 6 | 20.0 (NC ; NC) |  | - | - | - |
| D22 | 6 | 57.0 (3.86 ; 844) | 6 | 2.85 (0.193 ; 42.2) | 1/6 | 16.7 (0.4 ; 64.1) |
| D36 | 5 | 399 (NC ; NC) | 5 | 20.0 (NC ; NC) | 4/5 | 80.0 (28.4 ; 99.5) |

Participants in the control group received a primary series of two doses, on days 1 and 22, of CoV2 preS dTM-AS03 containing 10 ug D614 spike antigen

*Seroresponse (defined as ≥4-fold rise in neutralizing titer, post-/pre-vaccination)

CI, confidence interval; D, study day; GMT, geometric mean titer; GMTR, geometric mean titer ratio (geometric mean of individual titer ratios [D22/D1 or D36/D1]); M: number of participants with available data for the relevant endpoint; N: number of participants in PPAS; NC, not calculated

2-sided 95% CI for GMT is based on the Student t-distribution of logarithmic transformation of the individual titers. 2-sided exact 95% CI for percentage of participants with 4-fold rise is based on the Clopper-Pearson method.

**Table S4. P**seudovirus neutralizing antibody responses against B.1.351 and D614G following monovalent B.1.351 booster - PPAS

**S4.A. By priming vaccine and age group**

|  | **BNT162b2-primed** **(N=302)** | | **mRNA-1273-primed (N=102)** | | **ChAdOx1nCoV-19-primed (N=119)** | | **Ad26.CoV2.S-primed (N=92)** | | **CoV2 preS dTM-AS03 (D614)-primed  (N=119)** | |
| --- | --- | --- | --- | --- | --- | --- | --- | --- | --- | --- |
|  | **M or n/M** | **GMT, GMTR or % (95% CI)** | **M or n/M** | **GMT, GMTR or % (95% CI)** | **M or n/M** | **GMT, GMTR or % (95% CI)** | **M or n/M** | **GMT, GMTR or % (95% CI)** | **M or n/M** | **GMT, GMTR or % (95% CI)** |
| **B.1.351** |  |  |  |  |  |  |  |  |  |  |
| **18–55 years** |  |  |  |  |  |  |  |  |  |  |
| GMT D1 | 256 | 200 (157; 254) | 65 | 296 (200; 440) | 57 | 49.7 (32.7; 75.6) | 22 | 306 (114; 822) | 4 | 76.2 (NC; NC) |
| GMT D15 | 279 | 7172 (6363; 8083) | 68 | 9146 (6734; 12421) | 62 | 5643 (4468; 7128) | 21 | 7900 (5309; 11754) | 4 | 15509 (NC; NC) |
| D15/D1 | 256 | 35.4 (28.1; 44.6) | 65 | 29.8 (18.6; 47.7) | 57 | 108 (71.2; 163) | 21 | 27.9 (10.5; 74.3) | 4 | 203 (NC; NC) |
| ≥4-fold rise* | 211/256 | 82.4 (77.2; 86.9) | 52/65 | 80.0 (68.2; 88.9) | 55/57 | 96.5 (87.9; 99.6) | 15/21 | 71.4 (47.8; 88.7) | 4/4 | 100 (39.8; 100) |
| **≥ 56 years** |  |  |  |  |  |  |  |  |  |  |
| GMT D1 | 43 | 83.4 (49.7; 140) | 19 | 158 (71.7; 350) | 30 | 35.8 (23.0; 55.7) | 8 | 167 (47.0; 595) | 60 | 71.9 (47.2; 109) |
| GMT D15 | 46 | 6175 (4271; 8927) | 25 | 10324 (6738; 15818) | 32 | 3115 (1962; 4945) | 9 | 4964 (1412; 17450) | 68 | 13180 (9571; 18151) |
| D15/D1 | 43 | 71.2 (38.3; 132) | 19 | 72.0 (33.2; 156) | 30 | 89.5 (53.0; 151) | 8 | 23.3 (4.32; 125) | 60 | 179 (105; 305) |
| ≥4-fold rise* | 38/43 | 88.4 (74.9; 96.1) | 18/19 | 94.7 (74.0; 99.9) | 28/30 | 93.3 (77.9; 99.2) | 7/8 | 87.5 (47.3; 99.7) | 54/60 | 90.0 (79.5; 96.2) |
| **D614G** |  |  |  |  |  |  |  |  |  |  |
| **18–55 years** |  |  |  |  |  |  |  |  |  |  |
| GMT D1 | 274 | 739 (599 ; 913) | 68 | 1331 (917 ; 1931) | 55 | 165 (105 ; 258) | 22 | 741 (267 ; 2059) | 4 | 189 (NC ; NC) |
| GMT D15 | 279 | 10165 (9082; 11377) | 68 | 13189 (9836; 17684) | 62 | 6817 (5453; 8521) | 22 | 11424 (7948; 16419) | 4 | 37633 (NC; NC) |
| GMTR D15/D1 | 274 | 13.9 (11.3; 17.1) | 68 | 9.91 (6.92; 14.2) | 55 | 41.0 (27.0; 62.2) | 22 | 15.4 (6.18; 38.5) | 4 | 199 (NC; NC) |
| ≥4-fold rise* | 198/274 | 72.3 (66.6; 77.5) | 47/68 | 69.1 (56.7; 79.8) | 49/55 | 89.1 (77.8; 95.9) | 14/22 | 63.6 (40.7; 82.8) | 4/4 | 100 (39.8; 100) |
| **≥ 56 years** |  |  |  |  |  |  |  |  |  |  |
| GMT D1 | 41 | 298 (164 ; 541) | 24 | 861 (500 ; 1483) | 32 | 105 (61.4 ; 179) | 9 | 1456 (319 ; 6639) | 60 | 162 (95.9 ; 275) |
| GMT D15 | 46 | 9022 (6404; 12709) | 25 | 17545 (12042; 25562) | 32 | 3700 (2300; 5953) | 9 | 10046 (3184; 31699) | 68 | 24407 (17705; 33647) |
| GMTR D15/D1 | 41 | 29.5 (15.7; 55.3) | 24 | 20.2 (10.6; 38.4) | 32 | 35.3 (21.0; 59.2) | 9 | 6.90 (2.36; 20.2) | 60 | 146 (81.7; 259) |
| ≥4-fold rise* | 34/41 | 82.9 (67.9; 92.8) | 20/24 | 83.3 (62.6; 95.3) | 29/32 | 90.6 (75.0; 98.0) | 4/9 | 44.4 (13.7; 78.8) | 54/60 | 90.0 (79.5; 96.2) |

**S4.B. By priming platform and age group**

|  | **mRNA-primed** **(N=418)** | | **Adenovirus-vectored vaccine-primed (N=125)** | |
| --- | --- | --- | --- | --- |
|  | **M or n/M** | **GMT, GMTR or % (95% CI)** | **M or n/M** | **GMT, GMTR or % (95% CI)** |
| **B.1.351** |  |  |  |  |
| **18–55 years** |  |  |  |  |
| GMT D1 | 321 | 216 (176 ; 267) | 79 | 82.5 (53.3 ; 128) |
| GMT D15 | 347 | 7522 (6718; 8422) | 83 | 6145 (5032; 7503) |
| D15/D1 | 321 | 34.2 (27.8; 42.0) | 78 | 74.8 (49.6; 113) |
| ≥4-fold rise* | 263/321 | 81.9 (77.3; 86.0) | 70/78 | 89.7 (80.8; 95.5) |
| **≥ 56 years** |  |  |  |  |
| GMT D1 | 62 | 102 (66.2 ; 156) | 38 | 49.6 (31.4 ; 78.3) |
| GMT D15 | 71 | 7400 (5583; 9809) | 41 | 3450 (2247; 5298) |
| D15/D1 | 62 | 71.4 (44.3; 115) | 38 | 67.4 (39.6; 115) |
| ≥4-fold rise* | 56/62 | 90.3 (80.1; 96.4) | 35/38 | 92.1 (78.6; 98.3) |
| **D614G** |  |  |  |  |
| **18–55 years** |  |  |  |  |
| GMT D1 | 342 | 831 (690 ; 1001) | 77 | 253 (162 ; 396) |
| GMT D15 | 347 | 10697 (9611; 11907) | 84 | 7804 (6435; 9462) |
| GMTR D15/D1 | 342 | 13.0 (10.8; 15.6) | 77 | 31.0 (20.8; 46.1) |
| ≥4-fold rise* | 245/342 | 71.6 (66.5; 76.4) | 63/77 | 81.8 (71.4; 89.7) |
| **≥ 56 years** |  |  |  |  |
| GMT D1 | 65 | 441 (285 ; 681) | 41 | 187 (102 ; 343) |
| GMT D15 | 71 | 11402 (8757; 14846) | 41 | 4607 (2958; 7176) |
| GMTR D15/D1 | 65 | 25.6 (16.3; 40.3) | 41 | 24.7 (15.0; 40.4) |
| ≥4-fold rise* | 54/65 | 83.1 (71.7; 91.2) | 33/41 | 80.5 (65.1; 91.2) |

*Seroresponse (defined as ≥4-fold rise in neutralizing titer, post-/pre-vaccination)

CI, confidence interval; D, study day; GMT, geometric mean titer; GMTR (geometric mean titer ratio): geometric mean of individual titer ratios (post-vaccination/pre-vaccination); M: number of participants with available data for the relevant endpoint; N: number of participants in PPAS; NC, not calculated

2-sided 95% CI for GMT is based on the Student t-distribution of logarithmic transformation of the individual titers. 2-sided exact 95% CI for percentage of participants with 4-fold rise is based on the Clopper-Pearson method.

**Table S5.** B.1.351 and D614G pseudovirus neutralizing antibody responses following a bivalent D614+B.1.351 booster dose – PPAS

**S5.A. By prior priming vaccine and by age group**

|  | **BNT162b2-primed** **(N=335)** | | **mRNA-1273-primed (N=98)** | | **ChAdOx1nCoV-19-primed (N=94)** | | **Ad26.CoV2.S-primed (N=34)** | |
| --- | --- | --- | --- | --- | --- | --- | --- | --- |
|  | **M or n/M** | **GMT, GMTR or % (95% CI)** | **M or n/M** | **GMT, GMTR or % (95% CI)** | **M or n/M** | **GMT, GMTR or % (95% CI)** | **M or n/M** | **GMT, GMTR or % (95% CI)** |
| **B.1.351** |  |  |  |  |  |  |  |  |
| **18–55 years** |  |  |  |  |  |  |  |  |
| GMT D1 | 249 | 151 (119; 191) | 62 | 300 (195; 463) | 57 | 39.3 (28.5; 54.1) | 23 | 138 (53.7; 356) |
| GMT D15 | 276 | 5087 (4511; 5737) | 66 | 6698 (5349; 8388) | 57 | 4056 (2998; 5486) | 25 | 3951 (2459; 6350) |
| D15/D1 | 248 | 34.2 (27.2; 43.0) | 61 | 21.5 (13.8; 33.4) | 56 | 107 (70.5; 161) | 23 | 26.2 (12.4; 55.3) |
| ≥4-fold rise* | 205/248 | 82.7 (77.4; 87.2) | 48/61 | 78.7 (66.3; 88.1) | 54/56 | 96.4 (87.7; 99.6) | 18/23 | 78.3 (56.3; 92.5) |
| **≥56 years** |  |  |  |  |  |  |  |  |
| GMT D1 | 50 | 106 (57.9; 194) | 24 | 270 (112; 647) | 35 | 28.7 (20.5; 40.2) | 8 | 143 (13.4; 1521) |
| GMT D15 | 58 | 4100 (2940; 5716) | 29 | 6321 (3914; 10210) | 36 | 2025 (1276; 3212) | 9 | 10035 (5707; 17646) |
| D15/D1 | 50 | 38.2 (21.9; 66.6) | 24 | 22.8 (10.9; 47.5) | 35 | 71.8 (44.7; 115) | 8 | 68.9 (7.91; 599) |
| ≥4-fold rise* | 42/50 | 84.0 (70.9; 92.8) | 20/24 | 83.3 (62.6; 95.3) | 34/35 | 97.1 (85.1; 99.9) |  | 75.0 (34.9; 96.8) |
| **D614G** |  |  |  |  |  |  |  |  |
| **18–55 years** |  |  |  |  |  |  |  |  |
| GMT D1 | 270 | 585 (477 ; 718) | 64 | 1400 (945 ; 2074) | 56 | 138 (89.6 ; 213) | 24 | 510 (219 ; 1183) |
| GMT D15 | 276 | 8550 (7638; 9571) | 67 | 12496 (10155; 15376) | 57 | 6923 (5124; 9353) | 25 | 9160 (6152; 13637) |
| GMTR D15/D1 | 269 | 14.4 (11.8; 17.5) | 63 | 8.57 (5.98; 12.3) | 55 | 52.5 (33.0; 83.6) | 24 | 17.3 (8.96; 33.3) |
| ≥4-fold rise* | 203 | 75.5 (69.9; 80.5) | 39/63 | 61.9 (48.8; 73.9) | 49/55 | 89.1 (77.8; 95.9) | 19/24 | 79.2 (57.8; 92.9) |
| **≥56 years** |  |  |  |  |  |  |  |  |
| GMT D1 | 51 | 386 (208 ; 717) | 28 | 1206 (573 ; 2538) | 33 | 109 (65.4 ; 180) | 9 | 543 (74.1 ; 3975) |
| GMT D15 | 58 | 7883 (5747; 10812) | 29 | 12582 (8240; 19213) | 36 | 4492 (3201; 6303) | 9 | 22531 (10427; 48687) |
| GMTR D15/D1 | 51 | 23.1 (13.6; 39.4) | 28 | 11.1 (5.73; 21.3) | 33 | 44.9 (25.9; 77.9) | 9 | 41.5 (5.61; 307) |
| ≥4-fold rise* | 39/51 | 76.5 (62.5; 87.2) | 19/28 | 67.9 (47.6; 84.1) | 30/33 | 90.9 (75.7; 98.1) | 7/9 | 77.8 (40.0; 97.2) |

**S5.B. By priming platform and age group**

|  | **mRNA-primed** **(N=433)** | | **Adenovirus-vector primed (N=128)** | |
| --- | --- | --- | --- | --- |
|  | **M or n/M** | **GMT, GMTR or % (95% CI)** | **M or n/M** | **GMT, GMTR or % (95% CI)** |
| **B.1.351** |  |  |  |  |
| **18–55 years** |  |  |  |  |
| GMT D1 | 311 | 173 (140 ; 213) | 80 | 56.4 (39.1 ; 81.3) |
| GMT D15 | 342 | 5365 (4823; 5967) | 82 | 4024 (3136; 5163) |
| D15/D1 | 309 | 31.2 (25.4; 38.2) | 79 | 70.8 (48.2; 104) |
| ≥4-fold rise* | 253/309 | 81.9 (77.1; 86.0) | 72/79 | 91.1 (82.6; 96.4) |
| ≥**56 years** |  |  |  |  |
| GMT D1 | 74 | 143 (87.3 ; 236) | 43 | 38.7 (23.8 ; 63.0) |
| GMT D15 | 87 | 4736 (3611; 6211) | 45 | 2789 (1824; 4264) |
| D15/D1 | 74 | 32.3 (20.8; 50.1) | 43 | 71.2 (43.1; 118) |
| ≥4-fold rise* | 62/74 | 83.8 (73.4; 91.3) | 40/43 | 93.0 (80.9; 98.5) |
| **D614G** |  |  |  |  |
| **18–55 years** |  |  |  |  |
| GMT D1 | 334 | 691 (575 ; 832) | 80 | 204 (136 ; 307) |
| GMT D15 | 343 | 9208 (8330; 10179) | 82 | 7540 (5938; 9573) |
| GMTR D15/D1 | 332 | 13.0 (11.0; 15.5) | 79 | 37.5 (25.4; 55.3) |
| ≥4-fold rise* | 242/332 | 72.9 (67.8; 77.6) | 68/79 | 86.1 (76.5; 92.8) |
| ≥**56 years** |  |  |  |  |
| GMT D1 | 79 | 578 (356 ; 939) | 42 | 153 (86.6 ; 271) |
| GMT D15 | 87 | 9212 (7153; 11865) | 45 | 6201 (4343; 8855) |
| GMTR D15/D1 | 79 | 17.8 (11.8; 26.9) | 42 | 44.1 (25.2; 77.1) |
| ≥4-fold rise* | 58/79 | 73.4 (62.3; 82.7) | 37/42 | 88.1 (74.4; 96.0) |

*Seroresponse is defined as ≥4-fold rise in neutralizing titer, post-/pre-vaccination

CI, confidence interval; D, study day; GMT, geometric mean titer; GMTR (geometric mean titer ratio): geometric mean of individual titer ratios (post-/-vaccination/pre-vaccination)); M: number of participants with available data for the relevant endpoint; N: number of participants in PPAS

2-sided 95% CI for GMT is based on the Student t-distribution of logarithmic transformation of the individual titers. 2-sided exact 95% CI for percentage of participants with 4-fold rise is based on the Clopper-Pearson method.

.

**Table S6. Safety overview after any injection for booster groups by prior priming vaccine, overall and by age group (safety analysis set)**

|  | **Monovalent D614 N=803** | | | | | **Monovalent B.1.351 N=705** | | | | | **Bivalent D614+B.1.351 N=621** | | | | | |
| --- | --- | --- | --- | --- | --- | --- | --- | --- | --- | --- | --- | --- | --- | --- | --- | --- |
| **Participants experiencing at least one:** | **n/M** | **%** | **(95% CI)** | | | **n/M** | | **%** | | **(95% CI)** | **n/M** | | **%** | | **(95% CI)** | |
| **All ages** |  |  |  | | |  | |  | |  |  | |  | |  | |
| Immediate unsolicited AE | 0/803 | 0 | (0 ; 0.5) | | | 0/705 | | 0 | | (0 ; 0.5) | 0/621 | | 0 | | (0 ; 0.6) | |
| Immediate unsolicited AR | 0/803 | 0 | (0 ; 0.5) | | | 0/705 | | 0 | | (0 ; 0.5) | 0/621 | | 0 | | (0 ; 0.6) | |
| Solicited reaction | 682/799 | 85.4 | (82.7 ; 87.7) | | | 575/693 | | 83.0 | | (80.0 ; 85.7) | 524/616 | | 85.1 | | (82.0 ; 87.8) | |
| Solicited injection site reaction | 634/799 | 79.3 | (76.4 ; 82.1) | | | 534/693 | | 77.1 | | (73.7 ; 80.1) | 483/616 | | 78.4 | | (74.9 ; 81.6) | |
| Grade 3 injection site reaction | 20/799 | 2.5 | (1.5 ; 3.8) | | | 23/693 | | 3.3 | | (2.1 ; 4.9) | 12/616 | | 1.9 | | (1.0 ; 3.4) | |
| Solicited systemic reaction | 490/799 | 61.3 | (57.9 ; 64.7) | | | 416/693 | | 60.0 | | (56.3 ; 63.7) | 399/616 | | 64.8 | | (60.9 ; 68.5) | |
| Grade 3 systemic reaction | 53/799 | 6.6 | (5.0 ; 8.6) | | | 48/693 | | 6.9 | | (5.2 ; 9.1) | 39/616 | | 6.3 | | (4.5 ; 8.6) | |
| Unsolicited AE | 173/803 | 21.5 | (18.7 ; 24.6) | | | 163/705 | | 23.1 | | (20.1 ; 26.4) | 154/621 | | 24.8 | | (21.4 ; 28.4) | |
| Unsolicited AR | 71/803 | 8.8 | (7.0 ; 11.0) | | | 50/705 | | 7.1 | | (5.3 ; 9.2) | 49/621 | | 7.9 | | (5.9 ; 10.3) | |
| Unsolicited non-serious AE | 172/803 | 21.4 | (18.6 ; 24.4) | | | 161/705 | | 22.8 | | (19.8 ; 26.1) | 152/621 | | 24.5 | | (21.1 ; 28.1) | |
| Unsolicited non-serious AR | 71/803 | 8.8 | (7.0 ; 11.0) | | | 50/705 | | 7.1 | | (5.3 ; 9.2) | 48/621 | | 7.7 | | (5.8 ; 10.1) | |
| Unsolicited non-serious injection site AR | 22/803 | 2.7 | (1.7 ; 4.1) | | | 13/705 | | 1.8 | | (1.0 ; 3.1) | 14/621 | | 2.3 | | (1.2 ; 3.8) | |
| Unsolicited non-serious systemic AE | 159/803 | 19.8 | (17.1 ; 22.7) | | | 157/705 | | 22.3 | | (19.2 ; 25.5) | 145/621 | | 23.3 | | (20.1 ; 26.9) | |
| Unsolicited non-serious systemic AR | 54/803 | 6.7 | (5.1 ; 8.7) | | | 42/705 | | 6.0 | | (4.3 ; 8.0) | 37/621 | | 6.0 | | (4.2 ; 8.1) | |
| During study period |  |  | |  | |  | |  | |  |  |  | | | |  |
| AE leading to study discontinuation | 0/803 | 0 | | (0 ; 0.5) | | 0/705 | | 0 | | (0 ; 0.5) | 0/621 | 0 | | | | (0 ; 0.6) |
| SAE | 8/803 | 1.0 | | (0.4 ; 2.0) | | 8/705 | | 1.1 | | (0.5 ; 2.2) | 9/621 | 1.4 | | | | (0.7 ; 2.7) |
| Related SAE | 0/803 | 0 | | (0 ; 0.5) | | 0/705 | | 0 | | (0 ; 0.5) | 1/621 | 0.2 | | | | (0 ; 0.9) |
| Death | 0/803 | 0 | | (0 ; 0.5) | | 0/705 | | 0 | | (0 ; 0.5) | 0/621 | 0 | | | | (0 ; 0.6) |
| AESI | 0/803 | 0 | | (0 ; 0.5) | | 1/705 | | 0.1 | | (0 ; 0.8) | 0/621 | 0 | | | | (0 ; 0.6) |
| Related AESI | 0/803 | 0 | | (0 ; 0.5) | | 0/705 | | 0 | | (0 ; 0.5) | 0/621 | 0 | | | | (0 ; 0.6) |
| MAAE | 90/803 | 11.2 | | (9.1 ; 13.6) | | 89/705 | | 12.6 | | (10.3 ; 15.3) | 85/621 | 13.7 | | | | (11.1 ; 16.6) |
| Related MAAE | 2/803 | 0.2 | | (0 ; 0.9) | | 3/705 | | 0.4 | | (0.1 ; 1.2) | 1/621 | 0.2 | | | | (0 ; 0.9) |
| **18-55 years** |  |  | | | |  |  | | | |  | |  | | | |
| Immediate unsolicited AE | 0/502 | 0 | | | (0 ; 0.7) | 0/492 | 0 | | (0 ; 0.7) | | 0/483 | | 0 | (0 ; 0.8) | | |
| Immediate unsolicited AR | 0/502 | 0 | | | (0 ; 0.7) | 0/492 | 0 | | (0 ; 0.7) | | 0/483 | | 0 | (0 ; 0.8) | | |
| Any solicited reaction | 446/498 | 89.6 | | | (86.5 ; 92.1) | 425/484 | 87.8 | | (84.6 ; 90.6) | | 427/478 | | 89.3 | (86.2 ; 92.0) | | |
| Any solicited injection site reaction | 426/498 | 85.5 | | | (82.1 ; 88.5) | 402/484 | 83.1 | | (79.4 ; 86.3) | | 403/478 | | 84.3 | (80.7 ; 87.5) | | |
| Grade 3 solicited injection site reaction | 13/498 | 2.6 | | | (1.4 ; 4.4) | 14/484 | 2.9 | | (1.6 ; 4.8) | | 11/478 | | 2.3 | (1.2 ; 4.1) | | |
| Any solicited systemic AE | 331/498 | 66.5 | | | (62.1 ; 70.6) | 320/484 | 66.1 | | (61.7 ; 70.3) | | 331/478 | | 69.2 | (64.9 ; 73.4) | | |
| Grade 3 solicited systemic AE | 36/498 | 7.2 | | | (5.1 ; 9.9) | 37/484 | 7.6 | | (5.4 ; 10.4) | | 35/478 | | 7.3 | (5.2 ; 10.0) | | |
| Unsolicited AE | 118/502 | 23.5 | | | (19.9 ; 27.5) | 134/492 | 27.2 | | (23.3 ; 31.4) | | 129/483 | | 26.7 | (22.8 ; 30.9) | | |
| Unsolicited AR | 49/502 | 9.8 | | | (7.3 ; 12.7) | 48/492 | 9.8 | | (7.3 ; 12.7) | | 43/483 | | 8.9 | (6.5 ; 11.8) | | |
| Unsolicited non-serious AE | 117/502 | 23.3 | | | (19.7 ; 27.3) | 133/492 | 27.0 | | (23.2 ; 31.2) | | 127/483 | | 26.3 | (22.4 ; 30.5) | | |
| Unsolicited non-serious AR | 49/502 | 9.8 | | | (7.3 ; 12.7) | 48/492 | 9.8 | | (7.3 ; 12.7) | | 42/483 | | 8.7 | (6.3 ; 11.6) | | |
| Unsolicited non-serious injection site AR | 12/502 | 2.4 | | | (1.2 ; 4.1) | 13/492 | 2.6 | | (1.4 ; 4.5) | | 12/483 | | 2.5 | (1.3 ; 4.3) | | |
| Unsolicited non-serious systemic AE | 110/502 | 21.9 | | | (18.4 ; 25.8) | 129/492 | 26.2 | | (22.4 ; 30.3) | | 122/483 | | 25.3 | (21.4 ; 29.4) | | |
| Unsolicited non-serious systemic AR | 40/502 | 8.0 | | | (5.8 ; 10.7) | 40/492 | 8.1 | | (5.9 ; 10.9) | | 33/483 | | 6.8 | (4.7 ; 9.5) | | |
| During study period |  |  | | |  |  |  | |  | |  | |  |  | | |
| AE leading to study discontinuation | 0/502 | 0 | | | (0 ; 0.7) | 0/492 | 0 | | (0 ; 0.7) | | 0/483 | | 0 | (0 ; 0.8) | | |
| SAE | 4/502 | 0.8 | | | (0.2 ; 2.0) | 3/492 | 0.6 | | (0.1 ; 1.8) | | 7/483 | | 1.4 | (0.6 ; 3.0) | | |
| Related SAE | 0/502 | 0 | | | (0 ; 0.7) | 0/492 | 0 | | (0 ; 0.7) | | 1/483 | | 0.2 | (0 ; 1.1) | | |
| Death | 0/502 | 0 | | | (0 ; 0.7) | 0/492 | 0 | | (0 ; 0.7) | | 0/483 | | 0 | (0 ; 0.8) | | |
| AESI | 0/502 | 0 | | | (0 ; 0.7) | 0/492 | 0 | | (0 ; 0.7) | | 0/483 | | 0 | (0 ; 0.8) | | |
| Related AESI | 0/502 | 0 | | | (0 ; 0.7) | 0/492 | 0 | | (0 ; 0.7) | | 0/483 | | 0 | (0 ; 0.8) | | |
| MAAE | 54/502 | 10.8 | | | (8.2 ; 13.8) | 57/492 | 11.6 | | (8.9 ; 14.7) | | 68/483 | | 14.1 | (11.1 ; 17.5) | | |
| Related MAAE | 1/502 | 0.2 | | | (0 ; 1.1) | 3/492 | 0.6 | | (0.1 ; 1.8) | | 1/483 | | 0.2 | (0 ; 1.1) | | |
| **≥56 years** |  |  | | | | | | | | |  | | | | | |
| Any immediate unsolicited AE within 30 minutes after injection | 0/301 | 0 | | | (0 ; 1.2) | 0/213 | 0 | | (0 ; 1.7) | | 0/138 | | 0 | (0 ; 2.6) | | |
| Related immediate unsolicited AE after injection | 0/301 | 0 | | | (0 ; 1.2) | 0/213 | 0 | | (0 ; 1.7) | | 0/138 | | 0 | (0 ; 2.6) | | |
| Any solicited reaction | 236/301 | 78.4 | | | (73.3 ; 82.9) | 150/209 | 71.8 | | (65.1 ; 77.8) | | 97/138 | | 70.3 | (61.9 ; 77.8) | | |
| Any solicited injection site reaction | 208/301 | 69.1 | | | (63.5 ; 74.3) | 132/209 | 63.2 | | (56.2 ; 69.7) | | 80/138 | | 58.0 | (49.3 ; 66.3) | | |
| Grade 3 solicited injection site reaction | 7/301 | 2.3 | | | (0.9 ; 4.7) | 9/209 | 4.3 | | (2.0 ; 8.0) | | 1/138 | | 0.7 | (0 ; 4.0) | | |
| Any solicited systemic AE | 159/301 | 52.8 | | | (47.0 ; 58.6) | 96/209 | 45.9 | | (39.0 ; 52.9) | | 68/138 | | 49.3 | (40.7 ; 57.9) | | |
| Grade 3 solicited systemic AE | 17/301 | 5.6 | | | (3.3 ; 8.9) | 11/209 | 5.3 | | (2.7 ; 9.2) | | 4/138 | | 2.9 | (0.8 ; 7.3) | | |
| Unsolicited AE | 55/301 | 18.3 | | | (14.1 ; 23.1) | 29/213 | 13.6 | | (9.3 ; 19.0) | | 25/138 | | 18.1 | (12.1 ; 25.6) | | |
| Unsolicited AR | 22/301 | 7.3 | | | (4.6 ; 10.9) | 2/213 | 0.9 | | (0.1 ; 3.4) | | 6/138 | | 4.3 | (1.6 ; 9.2) | | |
| Unsolicited non-serious AE | 55/301 | 18.3 | | | (14.1 ; 23.1) | 28/213 | 13.1 | | (8.9 ; 18.4) | | 25/138 | | 18.1 | (12.1 ; 25.6) | | |
| Unsolicited non-serious AR | 22/301 | 7.3 | | | (4.6 ; 10.9) | 2/213 | 0.9 | | (0.1 ; 3.4) | | 6/138 | | 4.3 | (1.6 ; 9.2) | | |
| Unsolicited non-serious injection site AR | 10/301 | 3.3 | | | (1.6 ; 6.0) | 0/213 | 0 | | (0 ; 1.7) | | 2/138 | | 1.4 | (0.2 ; 5.1) | | |
| Unsolicited non-serious systemic AE | 49/301 | 16.3 | | | (12.3 ; 20.9) | 28/213 | 13.1 | | (8.9 ; 18.4) | | 23/138 | | 16.7 | (10.9 ; 24.0) | | |
| Unsolicited non-serious systemic AR | 14/301 | 4.7 | | | (2.6 ; 7.7) | 2/213 | 0.9 | | (0.1 ; 3.4) | | 4/138 | | 2.9 | (0.8 ; 7.3) | | |
| During study period |  |  | | |  |  |  | |  | |  | |  |  | | |
| AE leading to study discontinuation | 0/301 | 0 | | | (0 ; 1.2) | 0/213 | 0 | | (0 ; 1.7) | | 0/138 | | 0 | (0 ; 2.6) | | |
| SAE | 4/301 | 1.3 | | | (0.4 ; 3.4) | 5/213 | 2.3 | | (0.8 ; 5.4) | | 2/138 | | 1.4 | (0.2 ; 5.1) | | |
| Related SAE | 0/301 | 0 | | | (0 ; 1.2) | 0/213 | 0 | | (0 ; 1.7) | | 0/138 | | 0 | (0 ; 2.6) | | |
| Death | 0/301 | 0 | | | (0 ; 1.2) | 0/213 | 0 | | (0 ; 1.7) | | 0/138 | | 0 | (0 ; 2.6) | | |
| AESI | 0/301 | 0 | | | (0 ; 1.2) | 1/213 | 0.5 | | (0 ; 2.6) | | 0/138 | | 0 | (0 ; 2.6) | | |
| Related AESI | 0/301 | 0 | | | (0 ; 1.2) | 0/213 | 0 | | (0 ; 1.7) | | 0/138 | | 0 | (0 ; 2.6) | | |
| MAAE | 36/301 | 12.0 | | | (8.5 ; 16.2) | 32/213 | 15.0 | | (10.5; 20.5) | | 17/138 | | 12.3 | (7.3 ; 19.0) | | |
| Related MAAE | 1/301 | 0.3 | | | (0 ; 1.8) | 0/213 | 0 | | (0 ; 1.7) | | 0/138 | | 0 | (0 ; 2.6) | | |

n: number of participants experiencing the endpoint; M: number of participants with available data for the relevant endpoint

*Related as assessed by the investigator

AE = adverse event. AESI = adverse event of special interest. AR = adverse reaction. n = number of participants experiencing the endpoint. M = number of participants with available data for the relevant endpoint. MAAE = medically-attended adverse event. pIMDs = potential immune-mediated diseases; SAE = serious adverse event. SafAS = Safety Analysis Set.

Related SAEs: reported by an investigator as related to the vaccine. SAEs for which information on the relationship was missing, were considered as related

**Table S7. Safety overview after any injection in the Control Group - SafAS**

|  | **Control (After Injection 1)   (N=473)** | | | | **Control (After Injection 2)  (N=445)** | | | | **Control (After Any Injection)  (N=473)** | | |  |
| --- | --- | --- | --- | --- | --- | --- | --- | --- | --- | --- | --- | --- |
| **Participants experiencing at least one:** | **n/M** | **%** | **(95% CI)** | **n** | **n/M** | **%** | **(95% CI)** | **n** | **n/M** | **%** | **(95% CI)** | **n** |
| Within 30 minutes |  |  |  |  |  |  |  |  |  |  |  |  |
| Immediate unsolicited AE | 5/473 | 1.1 | (0.3 ; 2.4) |  | 4/445 | 0.9 | (0.2 ; 2.3) |  | 9/473 | 1.9 | (0.9 ; 3.6) |  |
| Immediate unsolicited AR | 3/473 | 0.6 | (0.1 ; 1.8) |  | 4/445 | 0.9 | (0.2 ; 2.3) |  | 7/473 | 1.5 | (0.6 ; 3.0) |  |
| Within 7 days Solicited reaction | 378/459 | 82.4 | (78.6 ; 85.7) |  | 312/389 | 80.2 | (75.9 ; 84.1) |  | 409/460 | 88.9 | (85.7 ; 91.6) |  |
| Solicited injection site reaction | 344/459 | 74.9 | (70.7 ; 78.8) |  | 254/354 | 71.8 | (66.8 ; 76.4) |  | 375/460 | 81.5 | (77.7 ; 85.0) |  |
| Grade 3 injection site reaction | 18/459 | 3.9 | (2.3 ; 6.1) |  | 38/354 | 10.7 | (7.7 ; 14.4) |  | 51/460 | 11.1 | (8.4 ; 14.3) |  |
| Solicited systemic reaction | 291/456 | 63.8 | (59.2 ; 68.2) |  | 266/377 | 70.6 | (65.7 ; 75.1) |  | 359/460 | 78.0 | (74.0 ; 81.7) |  |
| Grade 3 systemic reaction | 56/456 | 12.3 | (9.4 ; 15.6) |  | 76/377 | 20.2 | (16.2 ; 24.6) |  | 115/460 | 25.0 | (21.1 ; 29.2) |  |
| Within 21 days |  |  |  |  |  |  |  |  |  |  |  |  |
| Unsolicited AE | 129/473 | 27.3 | (23.3 ; 31.5) |  | 107/445 | 24.0 | (20.1 ; 28.3) |  | 190/473 | 40.2 | (35.7 ; 44.7) |  |
| Unsolicited AR | 62/473 | 13.1 | (10.2 ; 16.5) |  | 72/445 | 16.2 | (12.9 ; 19.9) |  | 109/473 | 23.0 | (19.3 ; 27.1) |  |
| Unsolicited non-serious AE | 127/473 | 26.8 | (22.9 ; 31.1) |  | 106/445 | 23.8 | (19.9 ; 28.1) |  | 188/473 | 39.7 | (35.3 ; 44.3) |  |
| Unsolicited non-serious AR | 62/473 | 13.1 | (10.2 ; 16.5) |  | 72/445 | 16.2 | (12.9 ; 19.9) |  | 109/473 | 23.0 | (19.3 ; 27.1) |  |
| Unsolicited non-serious injection site AR | 13/473 | 2.7 | (1.5 ; 4.7) |  | 15/445 | 3.4 | (1.9 ; 5.5) |  | 26/473 | 5.5 | (3.6 ; 8.0) |  |
| Unsolicited non-serious systemic AE | 121/473 | 25.6 | (21.7 ; 29.8) |  | 96/445 | 21.6 | (17.8 ; 25.7) |  | 176/473 | 37.2 | (32.8 ; 41.7) |  |
| Unsolicited non-serious systemic AR | 51/473 | 10.8 | (8.1 ; 13.9) |  | 58/445 | 13.0 | (10.0 ; 16.5) |  | 87/473 | 18.4 | (15.0 ; 22.2) |  |
| During study period |  |  |  |  |  |  |  |  |  |  |  |  |
| AE leading to study discontinuation | 0/473 | 0 | (0 ; 0.8) |  | 1/445 | 0.2 | (0 ; 1.2) |  | 1/473 | 0.2 | (0 ; 1.2) |  |
| SAE | 3/473 | 0.6 | (0.1 ; 1.8) |  | 6/445 | 1.3 | (0.5 ; 2.9) |  | 9/473 | 1.9 | (0.9 ; 3.6) |  |
| Related SAE | 0/473 | 0 | (0 ; 0.8) |  | 0/445 | 0 | (0 ; 0.8) |  | 0/473 | 0 | (0 ; 0.8) |  |
| Death | 0/473 | 0 | (0 ; 0.8) |  | 0/445 | 0 | (0 ; 0.8) |  | 0/473 | 0 | (0 ; 0.8) |  |
| AESI | 0/473 | 0 | (0 ; 0.8) |  | 2/445 | 0.4 | (0.1 ; 1.6) |  | 2/473 | 0.4 | (0.1 ; 1.5) |  |
| Related AESI | 0/473 | 0 | (0 ; 0.8) |  | 1/445 | 0.2 | (0 ; 1.2) |  | 1/473 | 0.2 | (0 ; 1.2) |  |
| MAAE | 26/473 | 5.5 | (3.6 ; 8.0) | 37 | 35/445 | 7.9 | (5.5 ; 10.8) | 84 | 56/473 | 11.8 | (9.1 ; 15.1) | 121 |
| Related MAAE | 5/473 | 1.1 | (0.3 ; 2.4) | 6 | 4/445 | 0.9 | (0.2 ; 2.3) | 9 | 8/473 | 1.7 | (0.7 ; 3.3) | 15 |

n: number of participants experiencing the endpoint; M: number of participants with available data for the relevant endpoint; N: number of participants in SafAS for the Control Group; AR:

Reactions related to study vaccine

Related SAEs: causal relationship reported by an investigator as related. If the relationship is missing, the SAE will be considered as related.

The Control group received two monovalent D614 (10 µg CoV2 preS dTM-AS03 [D614]) primary vaccine doses given 21 days apart.

**Figure S1. Study design**

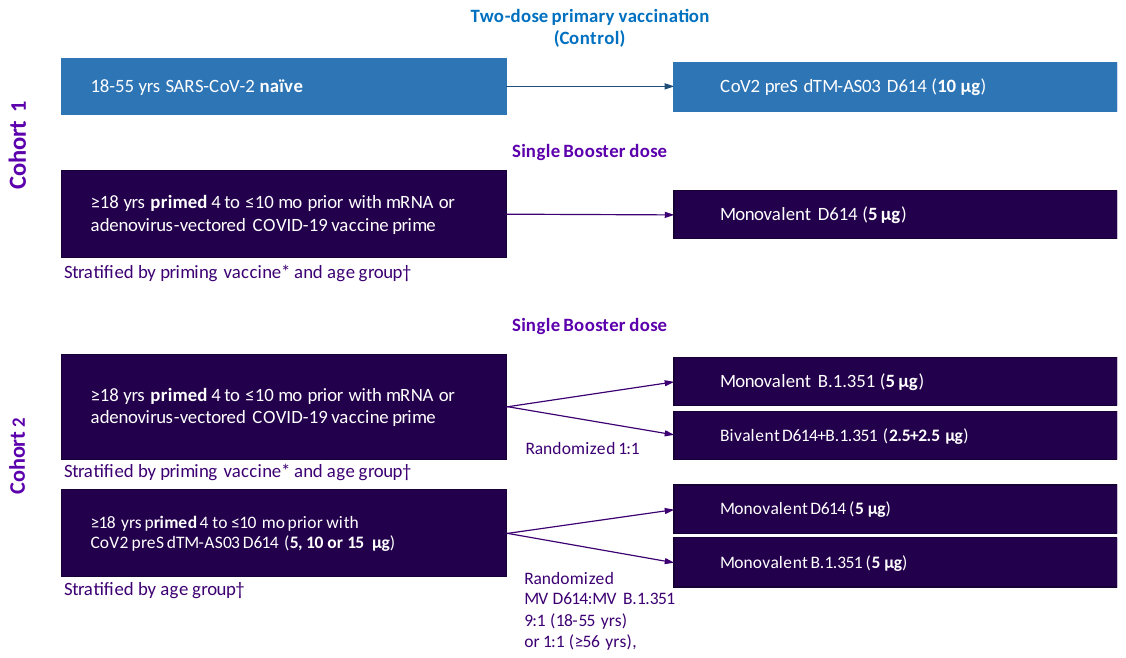

*mRNA COVID19 priming vaccines: BNT162b2 or mRNA-1273; adenovirus-vectored COVID19-priming vaccine: ChAdOx1nCoV-19 or Ad26.CoV2.S
†Age groups: 18–55 years and ≥56 years
mo, months; MV, monovalent

**Figure S2. Participant flow by CoV2 preS dTM-AS03 booster vaccine formulation**

D = study day. CMIAS = cell mediated immunity analysis set. FAS = full analysis set. SafAS = safety analysis set. PPAS = per protocol analysis set.
*Not due to an adverse event

**S2.A. CoV2 preS dTM-AS03 monovalent D614 booster**

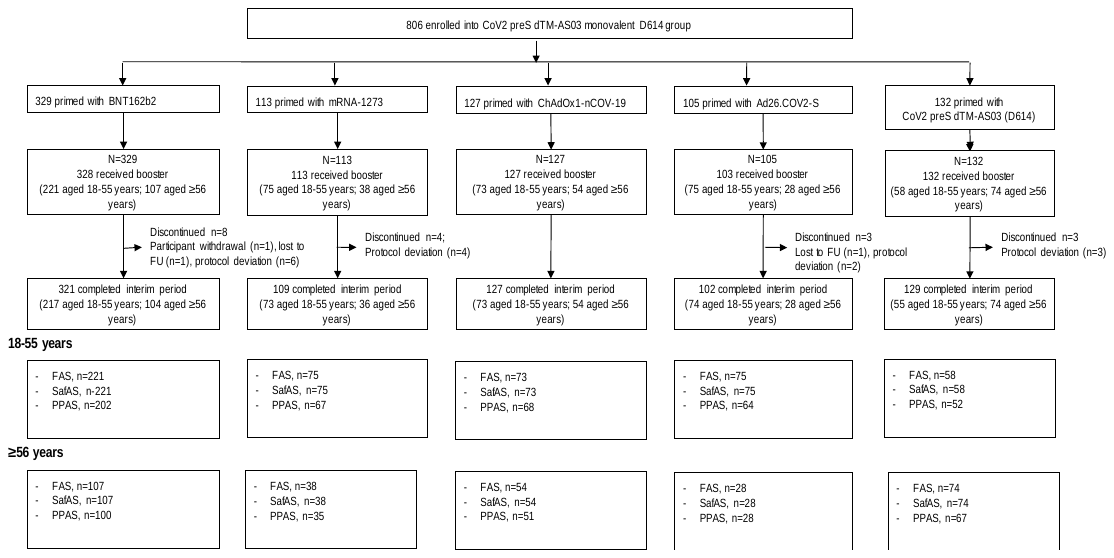

**S2.B. CoV2 preS dTM-AS03 monovalent B.1.351 booster**

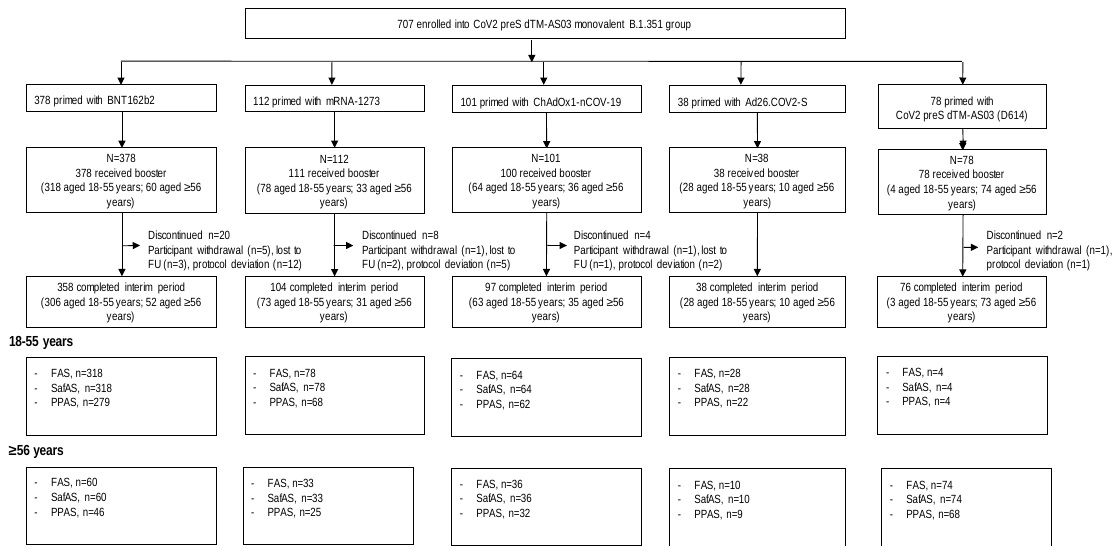

**S2.C. CoV2 preS dTM-AS03 bivalent D614+B.1.351 booster**

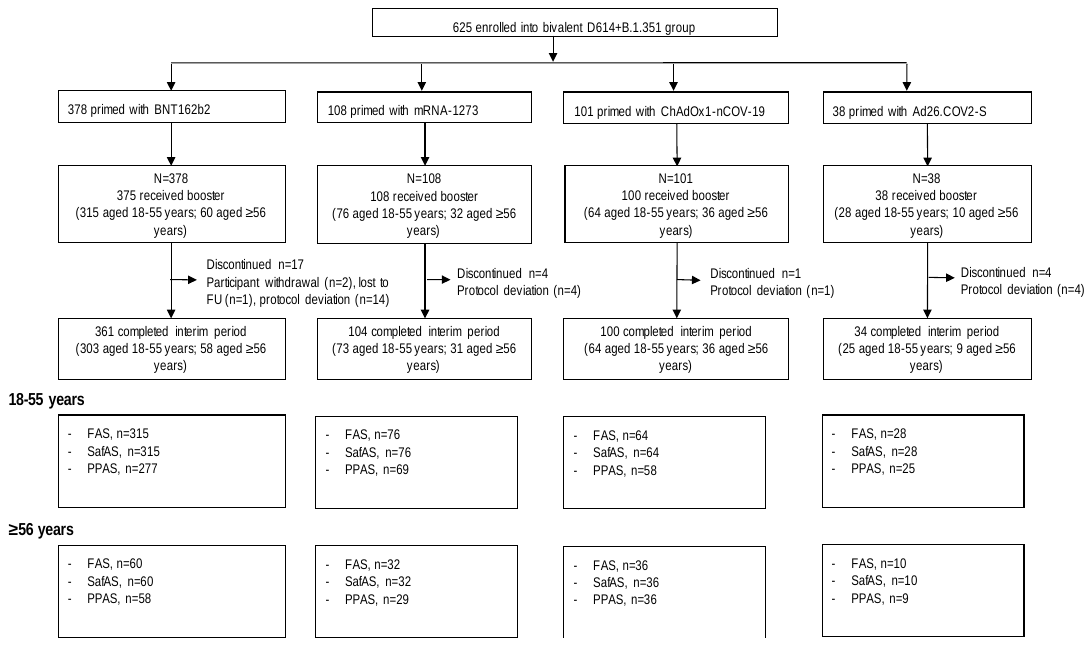

**S2.D. Control group**

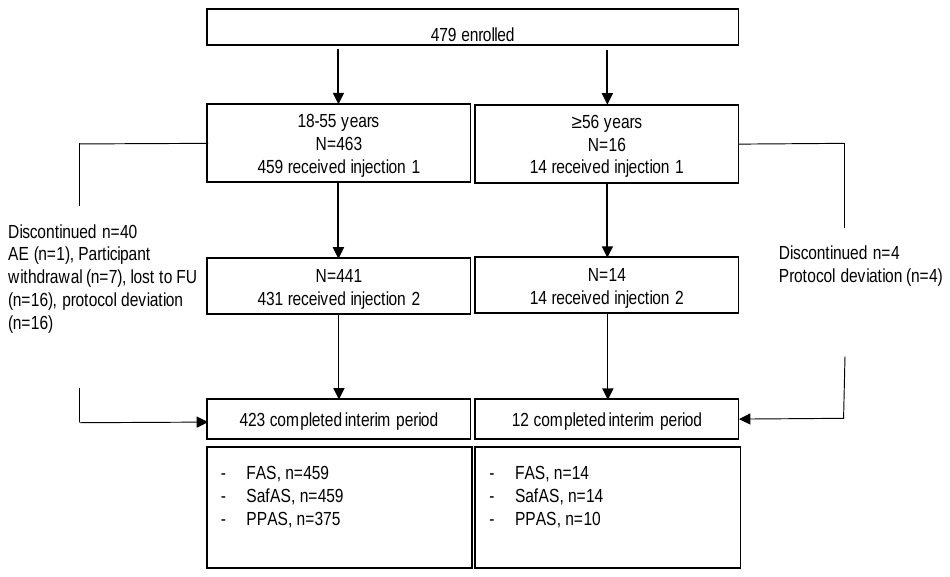

**Figure S3. Booster pseudovirus neutralizing antibody responses against D614G or B.1.351 strains for (A) monovalent D614, (B)** **monovalent B.1.351) and (C) bivalent D614+B.1.351 among ≥56 year olds, by prior priming vaccine – PPAS**

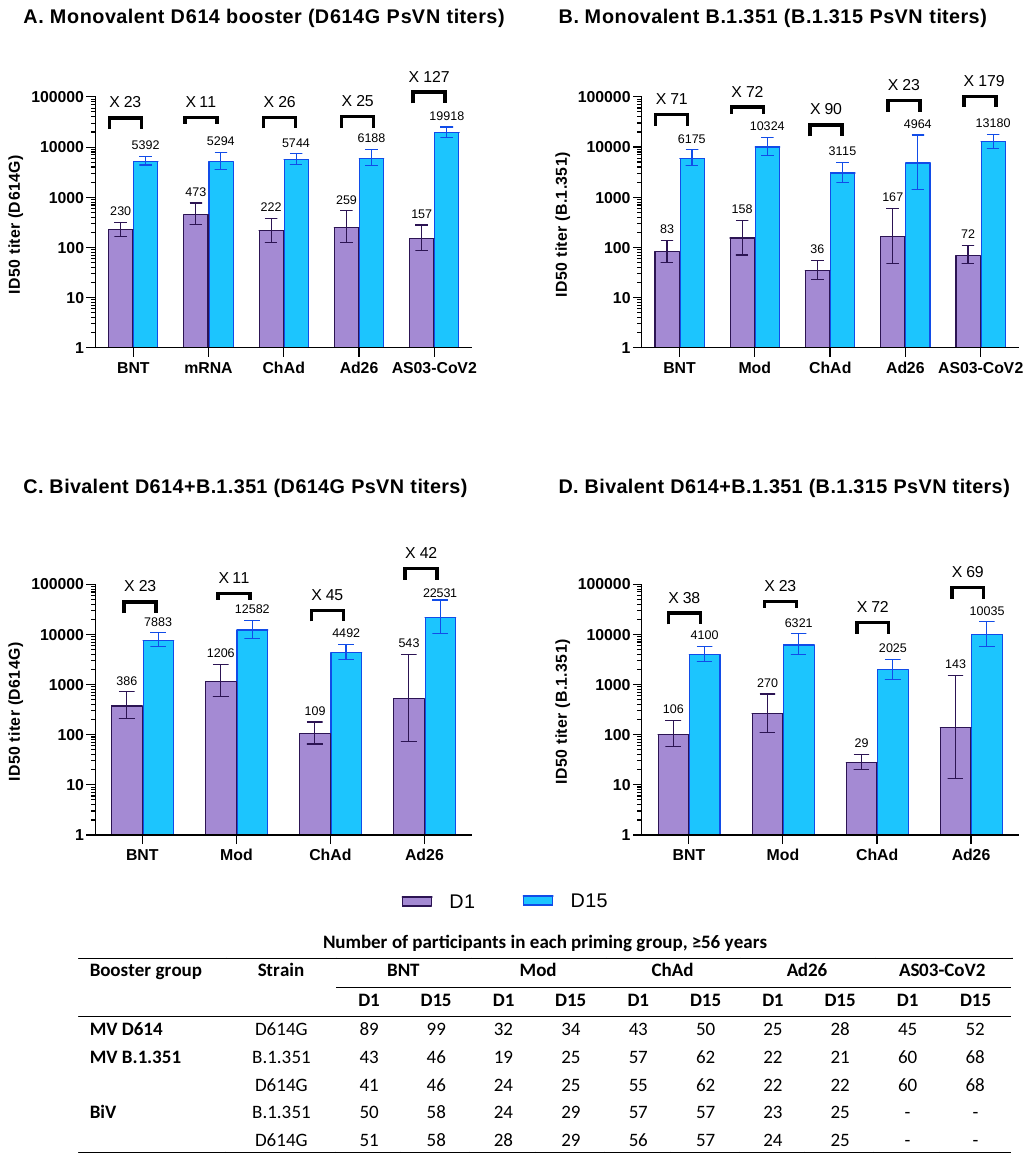
Graphs are annotated with geometric mean titers (above each bar) and geometric means of individual titer ratios (above grouped bars), calculated based on paired post-vaccination/pre-vaccination results. Error bars denote 95% CIs for the GMTs, calculated using normal approximation of log-transformed titers. Table shows number of participants with available data for at each timepoint.

D1, study day 1 (pre-booster); D15, study day 15 (14 days post-booster); BNT, BNT162b2-primed; Mod, mRNA-1273-primed; ChAd, ChAdOx1nCoV-19-primed; Ad26, Ad26.CoV2.S-primed; AS03-CoV2, CoV2 preS dTM-AS03 (D614)-primed; MV, monovalent; BiV, bivalent

**Figure S4. Solicited reactions by priming vaccine subgroup and by age group, for each booster group, SafAS**

**S4.A. Solicited injection site reactions within 7 days of injection**

*Monovalent D614 booster*

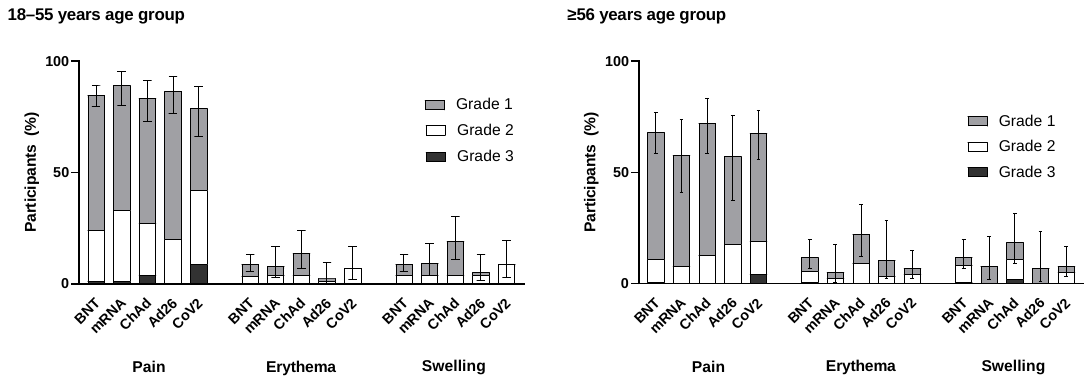

*Monovalent B.1.351 booster*

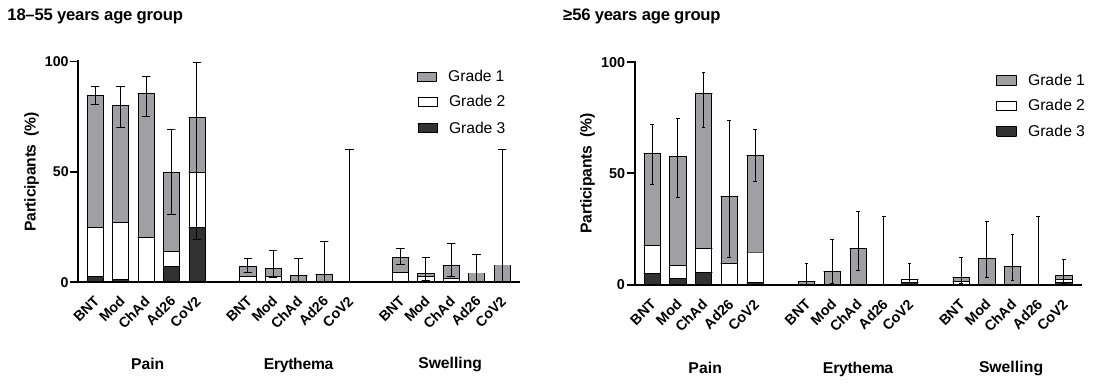

*Bivalent D614+B.1.351 booster*

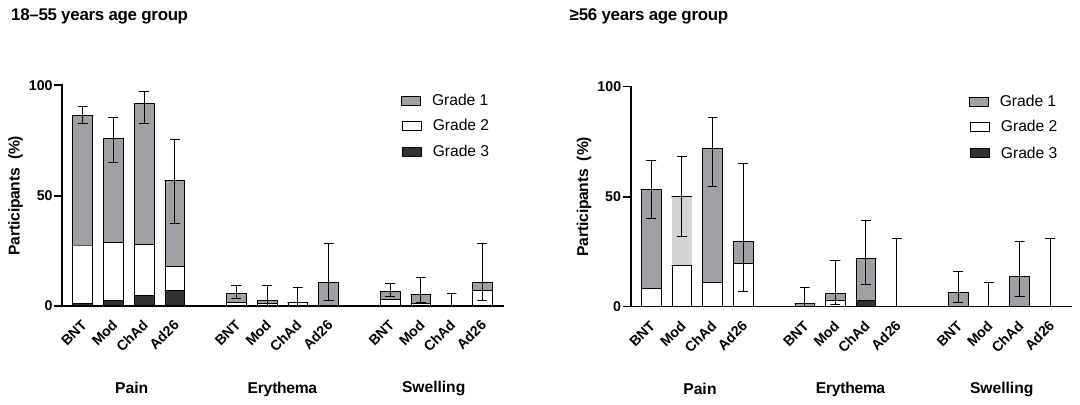

The percentages of participants experiencing at least one of the specified reactions are shown. Error bars denote 95% CIs for the percentages after any injection, calculated using the Clopper‑Pearson method.
BNT, BNT162b2-primed; Mod, mRNA-1273-primed; ChAd, ChAdOx1nCoV-19-primed; Ad26, Ad26.CoV2.S-primed; CoV2, CoV2 preS dTM-AS03 (D614)-primed

**S4.B. Solicited systemic reactions within 7 days of injection**

*Monovalent D614 booster*

18-55 years age group

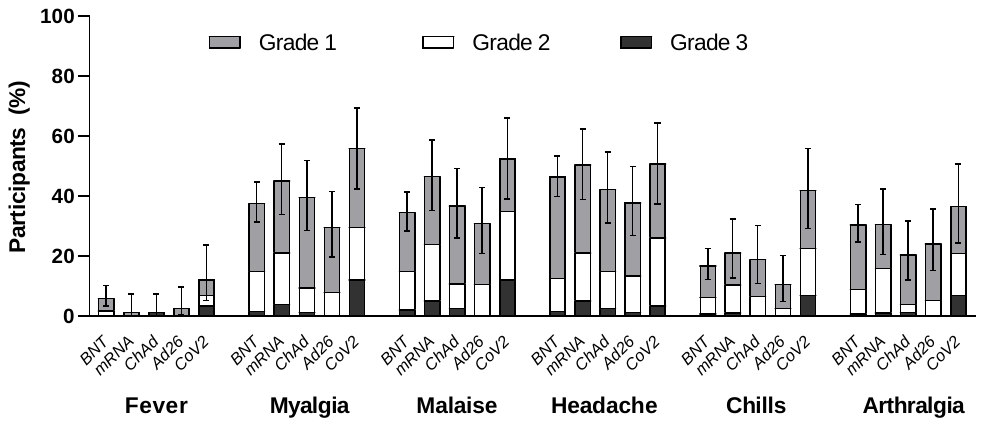

≥56 years age group

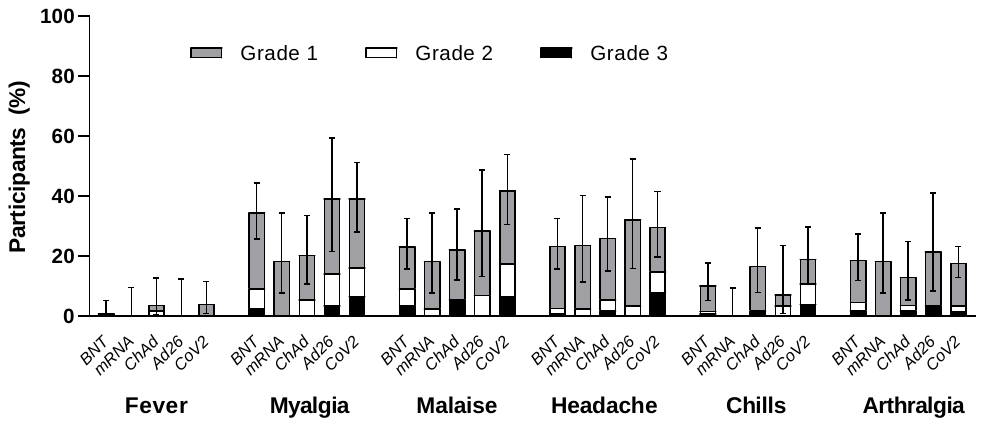

*Monovalent B.1.351 booster*

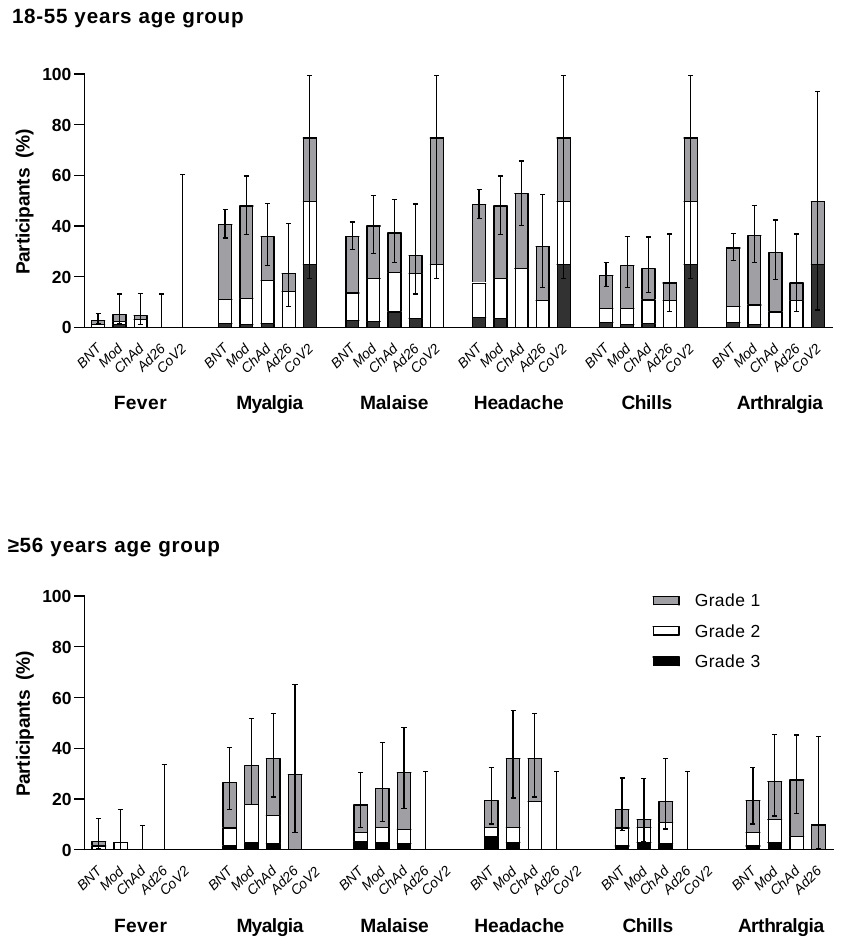

*Bivalent D614+B.1.351*

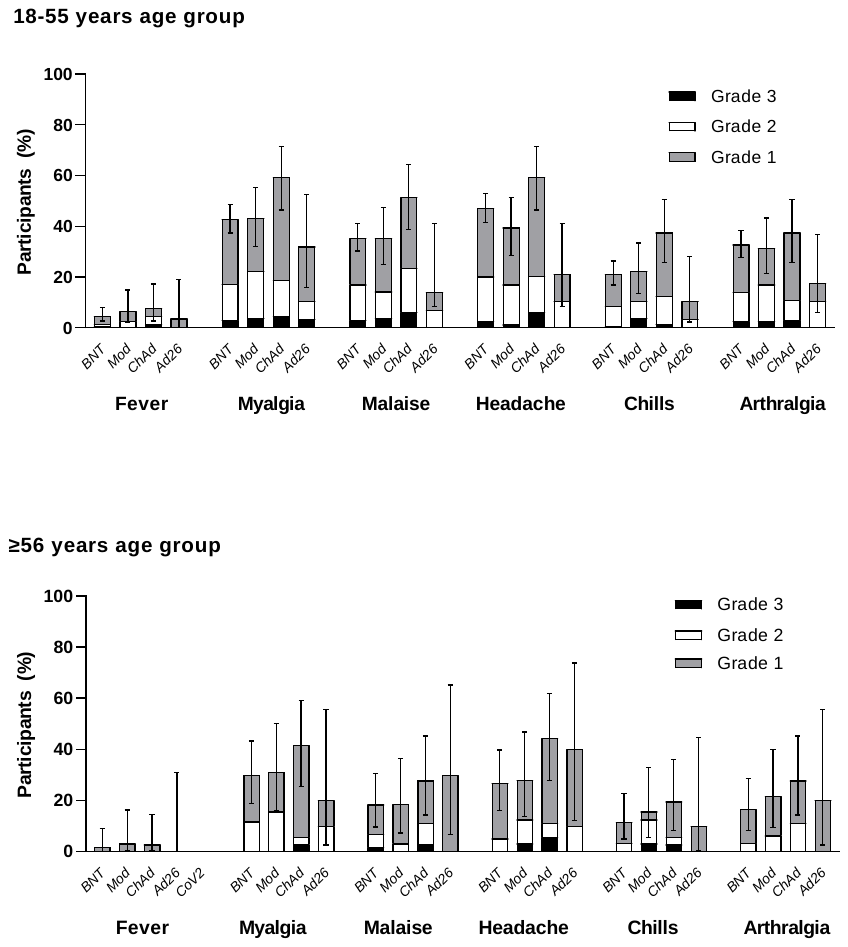

The percentages of participants experiencing at least one of the specified reactions are shown. Error bars denote 95% CIs for the percentages after any injection, calculated using the Clopper‑Pearson method.

BNT, BNT162b2-primed; mRNA, mRNA-1273-primed; ChAd, ChAdOx1nCoV-19-primed; Ad26, Ad26.CoV2.S-primed; CoV2, CoV2 preS dTM-AS03 (D614)-primed
